# Detection of Microsatellite Instability in Colorectal Cancer Tissue and Plasma samples using a new Multiplex Droplet Digital PCR kit

**DOI:** 10.1101/2024.04.12.24305349

**Authors:** Camille Evrard, Kariman Chaba, Justine Abdelli, Simon Garinet, Helene Blons, Aziz Zaanan, Jean-Baptiste Bachet, Leonor Benhaim, Claire Mulot, Audrey Didelot, Delphine Le Corre, Jerianne Lukban, Jennifer Yin, Adam Corner, Mai Ho, Valérie Taly, David Tougeron, Pierre Laurent-Puig

**Author notes:** Corresponding Authors: Pierre Laurent Puig and Valérie Taly, Centre de Recherche des Cordeliers, INSERM, Sorbonne Université, Université de Paris, Equipe Labellisée Ligue Nationale Contre le Cancer, CNRS SNC 5096, 15 Rue de L’école de Médecine, 75006 Paris, France. these authors contributed equally (Camille Evrard, Kariman Chaba).

## Abstract

Management of colorectal cancer (CRC) patients relies on the accurate determination of microsatellite instability (MSI) status. MSI status can have an influence on therapy decisions centered on immune checkpoint inhibitors. In this study a novel droplet digital PCR (ddPCR) kit for MSI status determination was validated across 2 separate CRC patient cohorts: 102 tumor samples from the ALGECOLS cohort and 129 plasma samples from the RASANC cohort. Each cohort was assessed for MSI status using the novel ddPCR kit and compared to historical and/or newly obtained results, (either immunohistochemistry analysis or PCR amplification). Concordance between ddPCR and conventional MSI determination methods for the analysis of tissue samples was 97.1% for ALGECOLS. When looking at positive ctDNA samples, a strong concordance was observed for the RASANC cohort. This study illustrates that ddPCR MSI testing represents a rapid, sensitive and accurate method with a strong correlation to established methods. Moreover, the ability of the described approach to monitor MSI status in both tumor and plasma enhances the potential for the use of MSI status in longitudinal monitoring of CRC patients.

## INTRODUCTION

With approximately 1.5 million new cases leading to 700.000 deaths each year, colorectal cancer (CRC) is a public health issue. The carcinogenesis of CRC is now well known with 75% of chromosomal instability, 25% of CpG island methylator phenotype (CIMP) and 15% of microsatellite instability (MSI) with an important overlap between CIMP and MSI (1). The determination of MSI status is important for the accurate management of CRC patients. In particular, the determination of mismatch repair status (i.e., proficient MMR (pMMR) or deficient MMR (dMMR) plays an important role in characterizing the sensitivity to immune checkpoint inhibitor with a recent approval of Pembrolizumab in dMMR/MSI metastatic CRC (mCRC) (2).

Microsatellites (MS) are repeated regions of 1 to 5 bases, widespread across the human genome. MSI is due to a defect in mismatch repair proteins (MSH2, MSH6, PMS2 and MLH1) resulting in either the increase or decrease of the number of repeats in microsatellite loci (3). Thus, MSI is a surrogate marker for deficient DNA mismatch repair. In the context of tumor biology, MSI is a marker of Lynch syndrome and a prognostic marker for CRC and other cancers such as endometrial cancers. Today only two techniques are validated to determine microsatellite/MMR status: immunohistochemistry (IHC) to detect the loss of MMR protein expression and molecular testing to assess fragment size variation of microsatellite loci (4). These techniques have some limitations including the fact that such experiments require significant quantities of DNA, are time-consuming and present limited sensitivity especially in the case of low tumor cell content within the sample. In addition, discordance between MSI and MMR status in tumor as well as heterogeneity of MSI status between primary tumor and metastases have been described (5,6).

Circulating tumor DNA (ctDNA) represents the fraction of circulating-free DNA (cfDNA) in blood originating from tumors that carry the same genetic alterations as those of the primary tumor (7). *KRAS* mutations were the first genetic alterations used to identify ctDNA in CRC patients (8) and ctDNA analysis is now used in routine clinical practice for determination of somatic mutations in tumors by a simple blood test called “liquid biopsy” (9). However, detection of MSI using ctDNA remains a challenge with very few case-series published up until now (10,11).

Droplet Digital PCR (ddPCR) is a well-established 3^rd^ generation nucleic acid amplification technique, where a fluorescent probe-based reaction is partitioned using emulsion technology into picolitre to nanolitre sized droplets (12,13). The droplet-partitioning process distributes target nucleic acid molecules evenly across the droplet population. Amplification of the target within the droplet population results in a fluorescent digital output within individual droplets. Each droplet develops fluorescent signal based on the presence or absence of the underlying target molecule and is subsequently individually detected and counted (13,14).

Droplet partitioning is particularly well suited for the detection of low-level nucleic acid fragments, such as small numbers of modified MS regions in a background of wild-type DNA. The use of ddPCR for MSI detection has several benefits over MSI analysis by traditional PCR followed by fragment length analysis or next generation sequencing (NGS) based approaches (15). These include improved sensitivity for low input amounts, improved sensitivity for low tumor content, the ability to detect ctDNA in blood samples, accurate quantification of MS allele frequency and total DNA, and reduced turnaround time for results (7,9,14).

The current research describes the use of a new Bio-Rad ddPCR MSI kit^®^, a 3 well, 5 markers ddPCR based method and its associated analysis software that utilizes an auto-calling algorithm to determine individual MSI status. This study describes the validation of the developed kit on both tumor and plasma samples from 2 independent CRC patient cohorts. These samples consisted of 102 tumor tissues (among which 96 frozen tissues) from the ALGECOLS multicentric prospective cohort (stage I-III CRC) and 129 plasma samples from the RASANC multicentric prospective cohort (all-comers mCRC) (9,16). The performance of the ddPCR MSI approach is compared to both traditional molecular analysis of MSI fragments and IHC of MMR proteins. Moreover, the software performance is assessed for tumor DNA (Fresh Frozen and Formalin-Fixed Paraffin-Embedded) and ctDNA.

## MATERIALS AND METHODS

Method description has been performed in order to comply with digital dMIQE2020 guidelines (Supplementary Table 1) (17).

### Samples

Fresh Frozen (FF) and Formalin-Fixed Paraffin-Embedded (FFPE) tumor DNA (n=102 including 96 FF and 6 FFPE tissue samples) and plasma DNA samples (n=129 DNA samples) from two patient cohorts were used in this work (see Figure 1 and Table 1 for summary for details of cohorts and samples).

**Figure 1.**
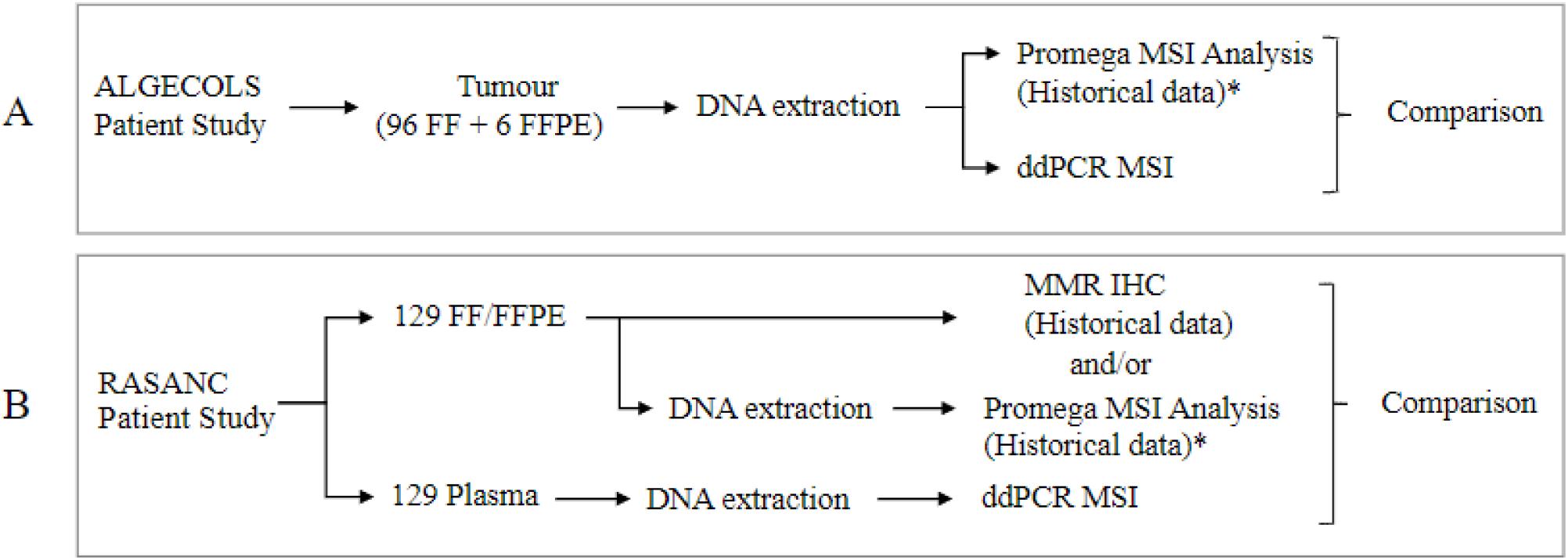
Summary of clinical testing and MSI analysis comparison for **(A)** ALGECOLS and **(B)** RASANC. (FF: Fresh Frozen, FFPE: formalin-fixed paraffin embedded, MSS: microsatellite stable, MSI-H: microsatellite instability-high, MMR IHC: mismatch repair immunohistochemistry). * The Promega MSI Analysis system used was the v1.2 for historical data and Promega Analysis System for repeats.

**Table 1.**
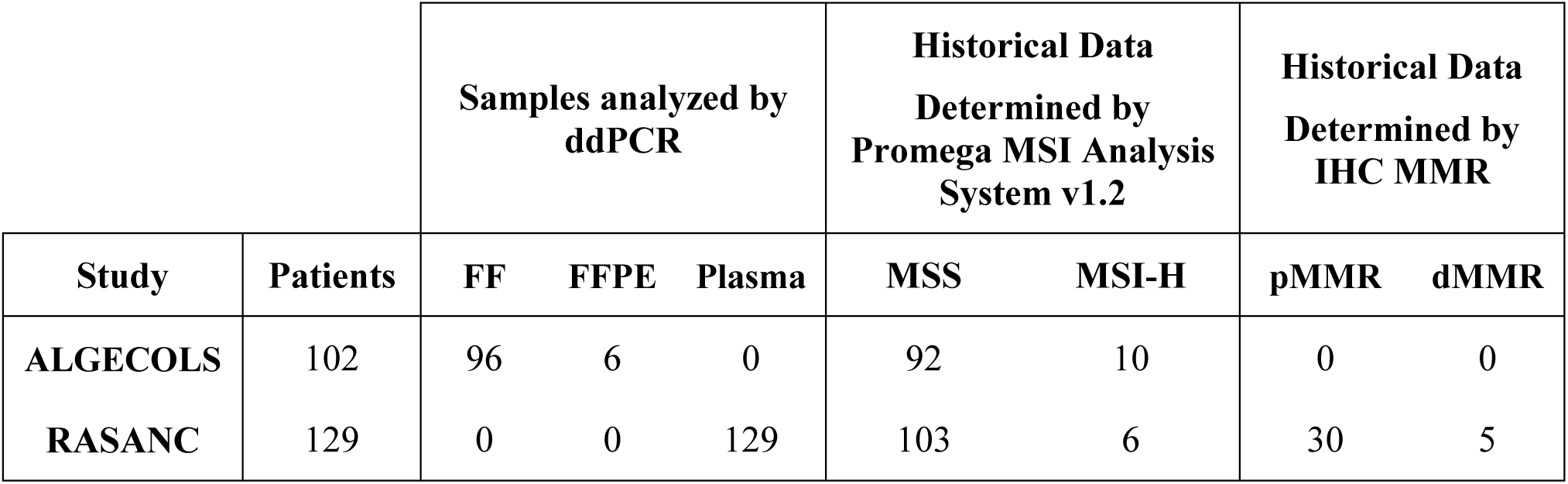
Summary of clinical CRC samples evaluated from ALGECOLS and RASANC studies. (FF: Fresh Frozen, FFPE: formalin-fixed paraffin embedded, MSS: microsatellite stable, MSI-H: microsatellite instability-high, pMMR: proficient mismatch repair, dMMR: deficient mismatch repair)

| Study | Patients | Samples analyzed by ddPCR |  |  | Historical Data Determined by Promega MSI Analysis System v1.2 |  | Historical Data Determined by IHC MMR |  |
| --- | --- | --- | --- | --- | --- | --- | --- | --- |
|  |  | FF | FFPE | Plasma | MSS | MSI-H | pMMR | dMMR |
| ALGECOLS | 102 | 96 | 6 | 0 | 92 | 10 | 0 | 0 |
| RASANC | 129 | 0 | 0 | 129 | 103 | 6 | 30 | 5 |

Cohort 1: Ninety-six (96) FF and 6 FFPE CRC samples with known MSI status were retrospectively selected from the ALGECOLS cohort (according to availability of samples) approved by the Ile-de-France ethics committee number 2 (NCT01198743) (16). The samples were collected from stage I to III patients. All samples were previously screened for microsatellite instability using the Promega MSI Analysis System v1.2^®^ (Promega Corporation, Madison, USA, cat. Number: MD1641) which targets 5 microsatellite markers (*BAT-25, BAT-26, NR-21, NR-24* and *Mono-27*), according to the manufacturer’s protocol (Promega Corporation). The absence of any mutant markers or the presence of 1 mutant marker was classified as microsatellite stable (MSS). The presence of 2 or more mutant markers was classified as microsatellite instability-high (MSI-H).

Cohort 2: From the RASANC protocol, approved by the Ile-de-France IV ethics committee, 129 cell-free DNA (cfDNA) samples were analyzed (according to availability) (9). The cfDNA was extracted from prospectively collected plasma samples from stage IV CRC patients. All mCRC patients were chemo-naïve at the time of collection. The cfDNA was extracted from 1 mL of plasma using the Promega Maxwell RSC cfDNA Plasma kit^®^ (Promega Corporation, Madison, USA, cat Number: AS1480) with an elution volume of 50 µL. MSI status was pre-determined for all patients by analyzing matched tumor collected from each patient using the Promega MSI Analysis System v1.2^®^ (n=92), IHC of four MMR proteins (n=20) or both (n=15). Data from tumor testing were collected retrospectively and only final records of MSI status (i.e., MSS or MSI-H) were available without individual marker status. No tumor samples were available for MSI ddPCR testing for this study.

### Sample Quantitation

All FF and FFPE DNA samples were quantified using the Qubit dsDNA BR Assay (Thermo Fisher Scientific, Waltham, USA, cat. Number: Q32853) and all cfDNA samples were quantified using the Qubit dsDNA HS Assay (Thermo Fisher Scientific, Waltham, USA, cat Number: Q32854). Afterward all FF and FFPE DNA samples were diluted in 10 mM Tris 0.1 mM EDTA buffer to ∼3 ng/µL.

### ddPCR Microsatellite Instability Test

Five mononucleotide microsatellites (*BAT-25, BAT-26, NR-21, NR-24* and *Mono-27*) were targeted to determine MSI status using the Bio-Rad ddPCR MSI RUO Kit (research use only, Bio-Rad Laboratories, Hercules, USA). The assay can detect indels as short as 1 bp by using a competitive probe drop-off design for all markers, where a set of two probes, one labeled with FAM and the other with HEX, compete for the same target sequence. When the target sequence has an alteration in repeat length, one probe is outcompeted correspondingly to the length change. For most patients, wild type DNA will be detected by both competing probes in approximately a fifty-fifty ratio. Since most patients are homozygous for each marker, the wild type will present predominantly as one cluster. In less common instances, where patients are heterozygous, wild type will present as two clusters with similar allele frequencies. Since most colorectal cancer patients with MSI-H express a mean deletion of ≥6 bp, thresholding is based on the 2-dimensional cluster location. With the aid of the clusters in the controls, the thresholding generally follows these guidelines: 1) droplets in the cluster with the lowest FAM and HEX signal plus the area approximately one-third the distance between other dominant clusters are negative, 2) droplets in any cluster orthogonal to the dominant negative cluster plus the area approximately one-third the distance from the wild type cluster and two-thirds the distance to the negative cluster are mutant, and 3) all remaining droplets are wild type.

All five markers are tested in a total of three reactions: MSI Assay 1 (*BAT-25* and *BAT-26*), MSI Assay 2 (*NR-21* and *NR-24*) and MSI Assay 3 (*Mono-27*). If 0 or 1 marker mutation is present, then the sample is classified as MSS. If 2 or more marker mutations are present, then the sample is classified as MSI-H. The three ddPCR MSI reactions were prepared in volumes of 22 µL, by combining 5.5 µL ddPCR Multiplex Supermix, 1.1 µL ddPCR MSI Assay 1, 2 or 3, 8.8 µL nuclease-free water and 6.6 µL of sample. For all DNA FF and FFPE samples, 2 units of uracil-DNA glycosylase (UDG) were added to the reaction mixture. All FF and FFPE DNA samples were tested in duplicate at an input of 18 ng per assay reaction, and 14 of these samples (7 MSS and 7 MSI-H) were also tested at an input of ∼1 ng and ∼5 ng per assay reaction. To determine the sensitivity of the technique, most cfDNA samples were tested neat due to limited sample, which resulted in a final input range of ∼0.3 to 12 ng per reaction. All cfDNA samples were tested separately from FF and FFPE DNA samples. All analyses were performed blind of MSI status.

For all ddPCR runs two controls were tested, a No Template Control (NTC) where water was added in place of sample and a Positive Control (PC) which consisted of wild type (microsatellite stable) and MSI-H plasmid DNA (one of each per target). For all ddPCR runs with cfDNA, a ddPCR thresholding control (TC) was also tested, which consisted of wild type and two MSI-H plasmid DNA or ultramer per target. Control reactions were prepared similarly to samples, and all control reactions were run in duplicate per assay.

After preparing the reaction, the mixture was then partitioned into nanolitre-sized droplets using a QX200 Droplet Generator (Bio-Rad Laboratories). The droplets were thermal cycled at an initial denaturation at 95 °C for 5 minutes, followed by 40 cycles of 94 °C for 30 seconds then 55 °C for 1 minute, followed by inactivation at 98 °C for 10 minutes and then a hold at 4 °C for ∼30 to 120 minutes on a C1000 Touch Thermal Cycler with 96-deep well (Bio-Rad Laboratories). The ramp rate for the thermal cycling was 2 °C/s. After thermal cycling droplets were then detected on a QX200 Droplet Reader using QuantaSoft v1.7 software (Bio-Rad Laboratories).

### ddPCR Microsatellite Instability Analysis

Data was analyzed using QuantaSoft Analysis Pro Software v1.0. For all FF and FFPE runs, the PC reaction wells were used to guide thresholding of droplet clusters. The mutant fractional abundance was calculated for each marker. It was used to determine instability taking into account the DNA damage observed in these sample types, which can affect signal amplitude and cause false positives. If the mutant fractional abundance exceeded 2.11%, 0.34%, 2.38%, 1.51%, and 0.56% for *BAT-25, BAT-26, NR-21, NR-24* and *Mono-27*, respectively, then the marker was considered positive (unstable) (Figure 2). These cut-offs were previously determined by Bio-Rad Laboratories by analyzing 135 CRC MSS FFPE samples and calculating the 95^th^ percentile of mutant fractional abundance (unpublished data). For all cfDNA runs, the PC reaction wells were used to guide thresholding of droplet clusters. If 4 or more mutant copies of a marker were detected, then the marker was considered positive (unstable). Cut-offs for cfDNA were previously determined by Bio-Rad Laboratories by analyzing 74 plasma samples from normal patients and calculating the 95^th^ percentile of mutant copies (unpublished data).

**Figure 2.**
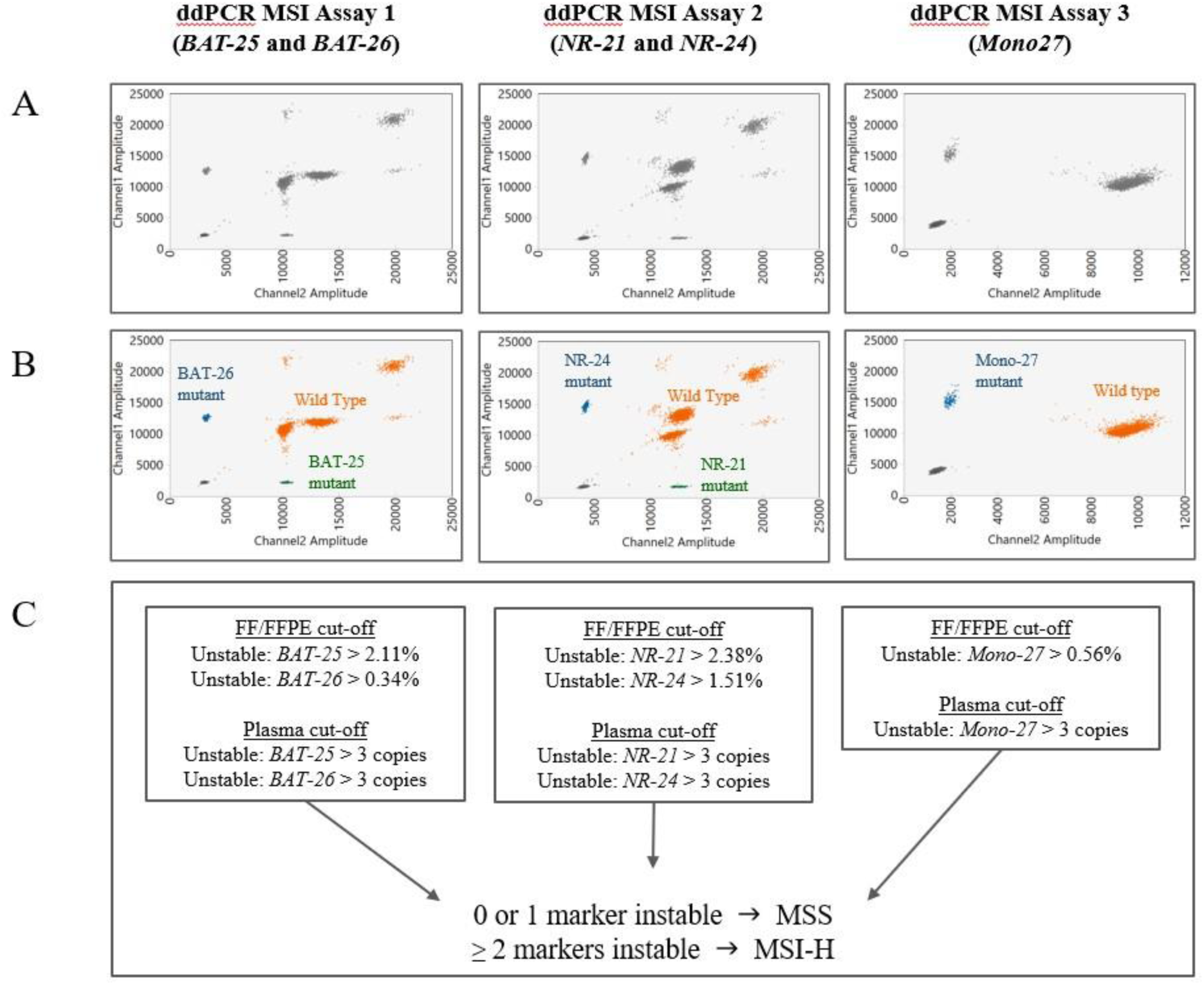
Bio-Rad ddPCR MSI assay analysis. **(A)** Unanalyzed 2-dimensional plots of Positive Control for MSI Assay 1, 2 and 3. **(B)** Thresholded 2-dimensional plots of Positive Control for MSI Assay 1, 2 and 3. **(C)** Determination of marker stability (*BAT-25, BAT-26, NR-21, NR-24* and *Mono-27*) and then MSI classification. Cut-offs for FF and FFPE samples are determined by mutant fractional abundance. Cut-offs for plasma sample are determined by mutant copies. (FF: Fresh Frozen, FFPE: formalin-fixed paraffin embedded, MSS: microsatellite stable, MSI-H: microsatellite instability-high, ddPCR: droplet digital PCR)

All FF, FFPE and cfDNA runs were also analyzed using an auto-thresholding algorithm. The algorithm works in two phases: a pre-trained neural network to generate initial droplet labels, followed by a droplet reclassification step using the on-plate PC wells. The neural network was developed by Bio-Rad Laboratories using a dataset of over 200 contrived and CRC patient FF, FFPE and cfDNA samples (more details in Supplementary Table 2). The auto-thresholding algorithm was applied using Bio-Rad QX Manager Premium Edition 2.0 software.

Reaction wells with a failure (i.e., unsuccessful droplet generation) or where droplet generation had less than 10,000 droplets were excluded from the analysis. A summary of all excluded data is in Supplementary Table 3.

All samples discordant between retrospective Promega MSI Analysis System v1.2 results and ddPCR MSI results were re-tested (see Supplementary Tables 4 and 5), if sample was available, using the Promega MSI Analysis System v1.2^®^ test according to the manufacturer’s protocol. The results of the repeat test were used for comparison to the ddPCR MSI results in tumors and plasma.

### Methylation-ddPCR assay testing of inconclusive or discordant samples

Plasma samples presenting with MSS status when matched tumor were classified MSI-H were tested by the Methylation-ddPCR assay (Met-ddPCR) as previously described to quantify the total cfDNA and the ctDNA fraction (9,16). Briefly, CRC methylation analysis of ccfDNA was performed using an assay designed for the simultaneous detection of *WIF1* and *NPY* gene hypermethylation. An albumin un-methylated sequence was used as a reference for measuring amplifiable DNA within each sample. Bisulfite conversion was performed on 20 ng of extracted DNA, whenever possible. For clinical samples, 20 µL of eluted DNA was bisulfite converted independently of sample concentration. The bisulfite reaction was carried out in a thermocycler at 98 °C for 12 minutes and 64 °C for 2 hours 35 minutes. The cleanup of bisulfite-converted DNA followed the recommendations of the manufacturer (EZ DNA Methylation-Gold, Zymo Research, Irvine, USA, cat. Number: D5006) and converted DNA was eluted in 10 µL of M-Elution Buffer and stored at -20 °C. Nine µL of converted DNA was used for each Met-ddPCR reaction. For each bisulfite conversion experiment, two different controls were included: a positive control (universal methylated human DNA standard, Zymo Research, Irvine, USA, cat. Number: D-5011-1) and a negative control (Human genomic DNA, Promega Corporation, Madison, USA, cat. Number: G304A). *BRAF V600E* ddPCR assay (*BRAF c.1799T>A)* was also performed as described previously (18). This ddPCR assay determines the concentration of target DNA in copies/µL, which are converted to ng/µL by applying an estimation of 303 copies per 1 ng of DNA as previously described (19).

## RESULTS

### Summary of the study

Detection of MS instability in CRC tumor tissue and plasma from two studies, ALGECOLS and RASANC were compared across two methods, Promega MSI Analysis system and ddPCR MSI (Figure 1). When available IHC MMR data were also used for the analysis.

### ddPCR MSI RUO Kit Sensitivity Evaluation

FF and FFPE samples from the ALGECOLS study were tested first to evaluate the sensitivity of the ddPCR MSI RUO kit. Fourteen of the 102 ALGECOLS samples, 7 MSS and 7 MSI-H were tested at three input levels: 18, 5 and 1 ng per reaction. Thirteen of the 14 samples at all three input levels showed MSI status concordant with the Promega MSI Analysis system v1.2 (Table 2). Only sample Alg-67, exhibited a MSS status by Promega analysis but a MSI-H status by ddPCR analysis at the highest input level of 18 ng per reaction with a mutant fractional abundance near the detection limit (i.e., <1.01% for all five markers). This data shows that ddPCR can accurately determine MSI-H status with as little as 1 ng of DNA.

**Table 2.**
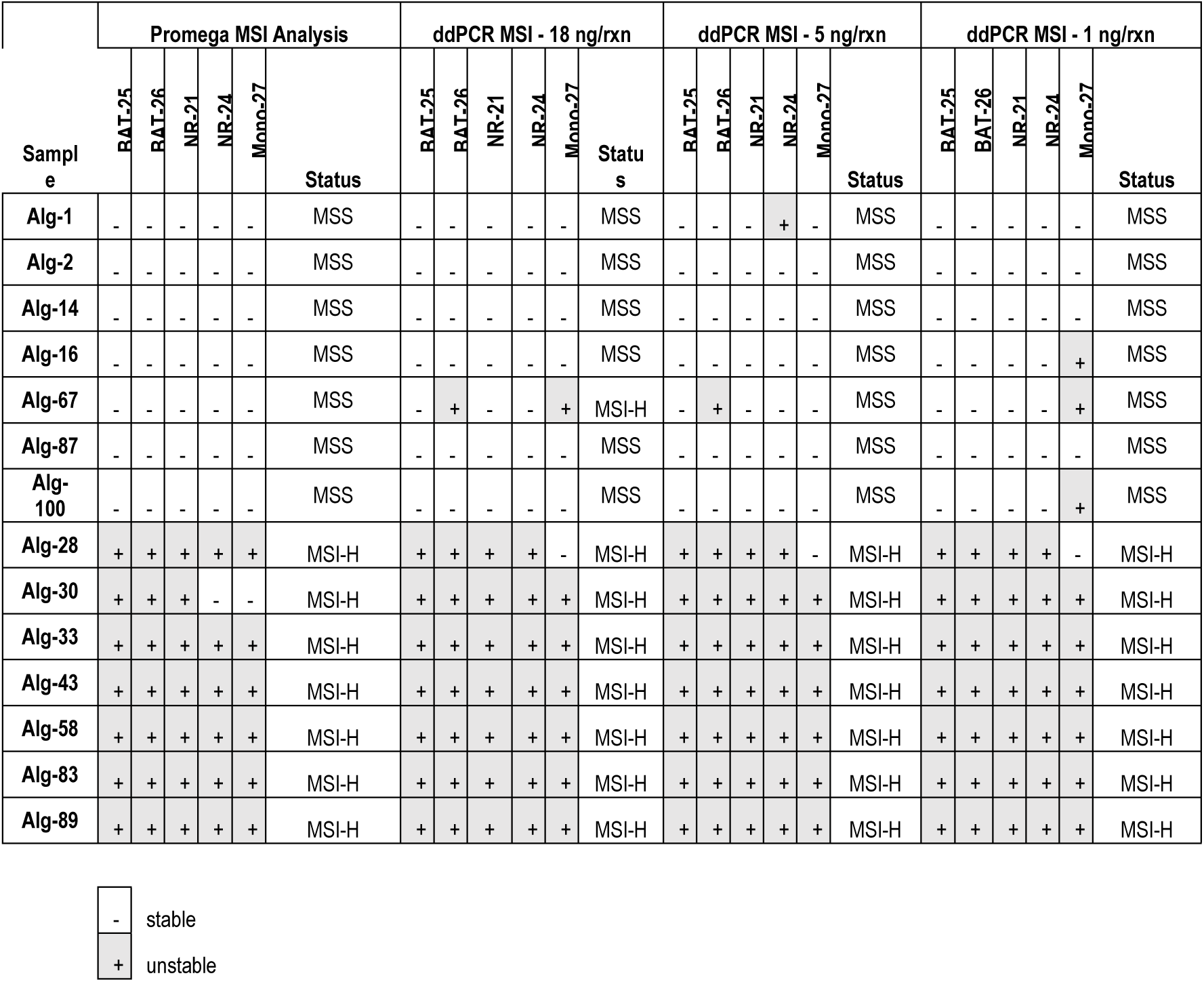
Comparison of ddPCR MSI with auto-thresholding for 14 FF and FFPE samples when tested at an input per well of 18, 5 and 1 ng/well, measured by Qubit dsDNA BR Assay. Seven (7) of the samples were determined as MSS and 7 as MSI-H by the MSI Analysis System. The ddPCR MSI results reflect data for replicate 1 of a total of 2 replicates. (MSS: microsatellite stable, MSI-H: microsatellite instability-high, ddPCR: droplet digital polymerase chain reaction)

|  |  |
| --- | --- |
| - | stable |
| + | unstable |

The ddPCR MSI results in Table 2 reflect data for replicate 1 of a total of 2 replicates. Data of both replicates were highly concordant (see Supplementary Table 6).

Additionally, the MSI status by ddPCR using a manual threshold (see Supplementary Table 7 and 8) were all concordant with the MSI status generated with the auto-thresholding algorithm, except for sample Alg-67 at an input of 18 ng per reaction (MSI-H with auto-thresholding only). As such, all the ddPCR MSI data presented in this article correspond to the auto-thresholded data of replicate 1 (See Supplementary Tables 7 and 8 for data corresponding to manual thresholding of replicate 1 and replicate 2, respectively).

### Analysis of tumor tissue samples from the ALGECOLS cohort

#### Concordance between ddPCR MSI and Promega MSI Analysis systems

The MSI status of the ALGECOLS tissues cohort (n=102, including the 14 described above) was previously tested using the Promega MSI Analysis System v1.2 resulting in 92 MSS and 10 MSI-H samples (historical data). Figures 3 show examples of 2-dimensional droplet plots for the three ddPCR MSI assay analyses of CRC FF samples that are either MSS or MSI-H, including a MSI-H sample showing detection of multiple alleles for all markers.

**Figure 3.**
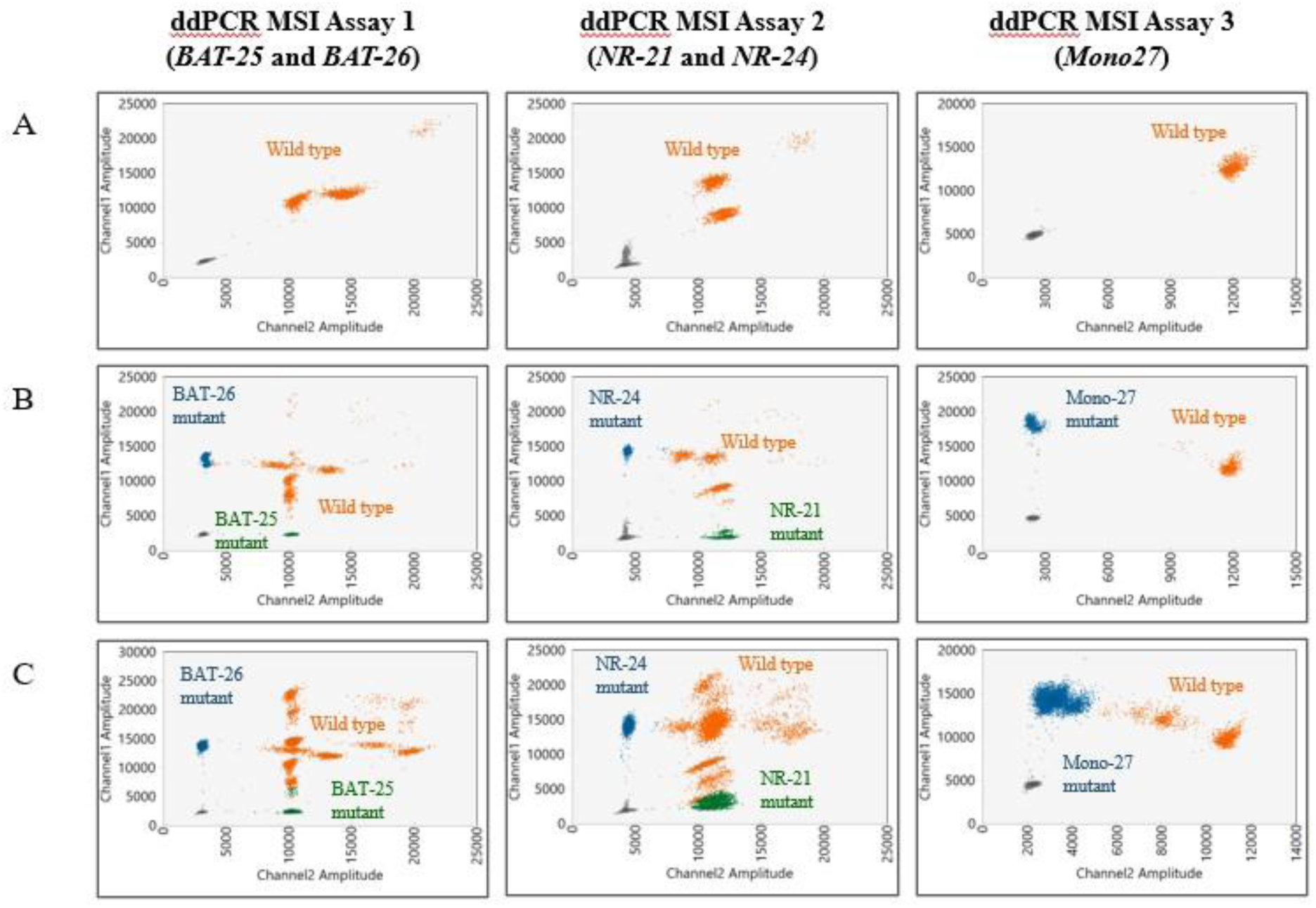
Example of auto-thresholded ddPCR MSI 2-dimensional droplet plots of CRC FF samples that are MSS (A, sample Alg-2), MSI-H (B, sample Alg-89) and MSI-H showing detection of several alleles for *BAT-25, NR-21, NR-24* and *Mono-27* markers (C, Alg-17).

After testing by the newly developed ddPCR MSI kit, the MSI status was concordant with the historical status (Promega MSI Analysis System) for 94 samples with 90 of them leading to concordant results for all 5 markers (83 MSS and 7 MSI-H). The 8 samples classified with discordant MSI status (see Supplementary Table 4), were re-tested using the Promega MSI Analysis System v1.2. Interestingly, 5 samples out of the 8 samples historically classified as MSS were re-classified as MSI-H leading to 87 MSS and 15 MSI-H (Table 3 and Supplementary Table 4). Those reclassified MSI-H FF or FFPE samples were called MSI-H by ddPCR. The 3 remaining discordant samples remained MSS by Promega Analysis System and MSI-H by ddPCR MSI kit analysis. Samples Alg-25 and Alg-67 showed low tumor content with low fractional abundances for all five markers. These results suggest that these samples were potentially misclassified by Promega MSI Analysis system due to a lack of sensitivity but properly classified MSI-H by the sensitive ddPCR MSI kit.

**Table 3.**
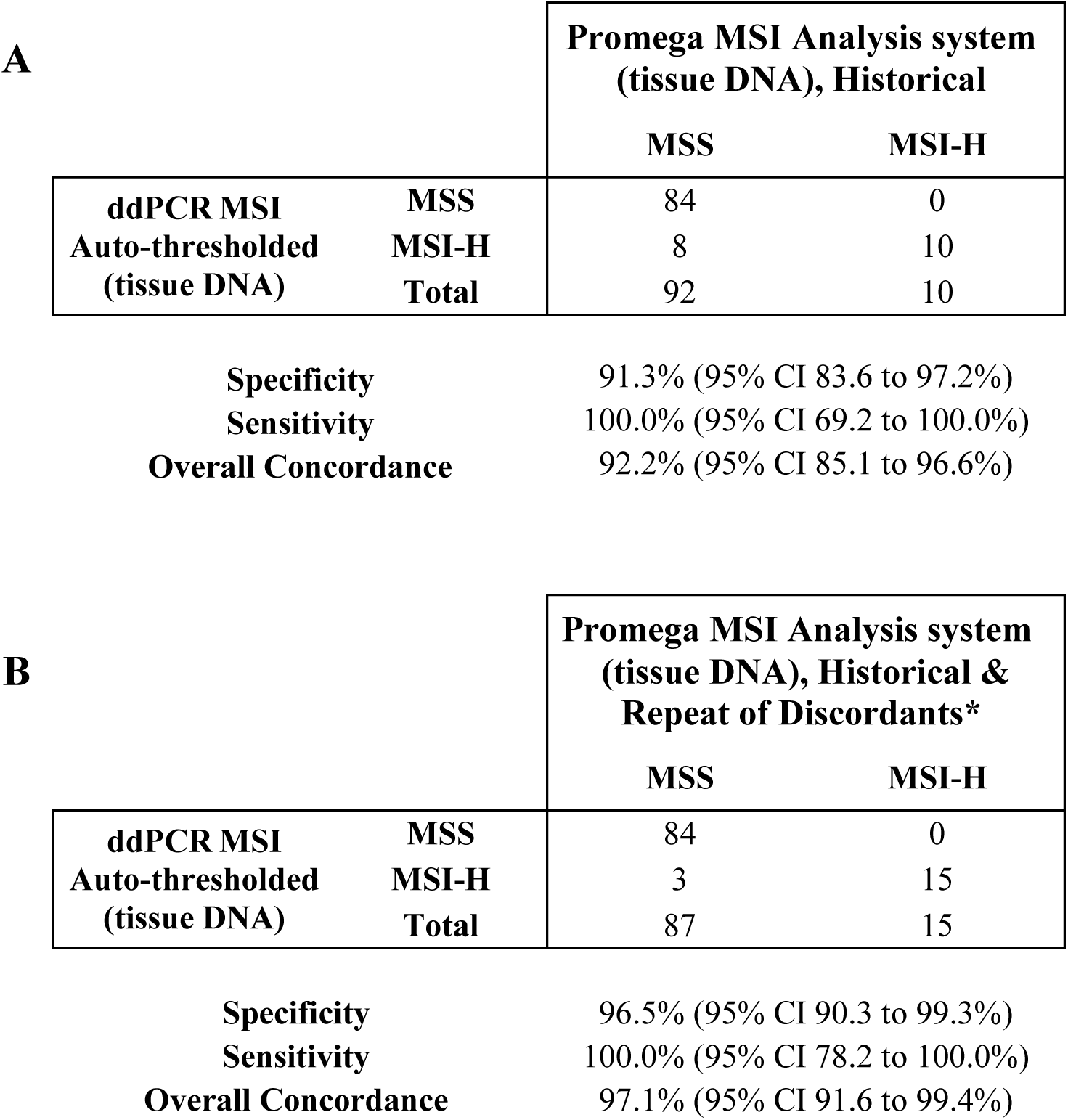
Concordance of FF and FFPE tissues from ALGECOLS cohort colorectal cancer patients between auto-thresholded ddPCR MSI with the historical Promega MSI Analysis System v1.2, historical results (A) and historical and repeat test of the discordants (B). (MSS: microsatellite stable, MSI-H: microsatellite instability-high, ddPCR: droplet digital polymerase chain reaction); * considered MSI status is the one obtained after retesting of discordant cases.

For the ALGECOLS cohort, intra- and inter-run reproducibility analysis of the ddPCR MSI assay are reported for the positive control and the samples in Supplementary Table 9 and 10, respectively. Intra- and inter-run reproducibility were consistently high.

### Analysis of the cfDNA samples from the RASANC cohort and comparison to tumor status

Of the 129 cfDNA samples tested from the RASANC study, 118 of the 120 samples of patients with MSS tumors by Promega analysis and 7 of the 9 samples of patients with MSI-H tumors by Promega analysis were concordant by ddPCR MSI analysis (Table 4). There were 4 discordant cases among 129 samples (3.1%). Figure 4 shows examples of 2-dimensional droplet plots for all three ddPCR MSI assays of CRC cfDNA samples that were classified as MSS (Figure 4A) or MSI-H (Figure 4B). Sample Ras-6 was unstable for *BAT-25 and NR-24* (low positive) by ddPCR MSI analysis. The repeat ddPCR MSI analysis re-classified this sample as MSS. The cfDNA and ctDNA content was then determined by Met-ddPCR analysis for this sample which demonstrated both high load of cfDNA and high fraction of ctDNA, which confirmed initial false positivity of this sample (Table 5). Sample Ras-39 was classified MSI-H by ddPCR analysis but MSS by Promega analysis. This sample also presented high load of cfDNA and ctDNA confirming this sample as discordant. The low fraction of MSI mutant DNA observed with ddPCR analysis is in favor of a heterogeneous tumor where the MSI-H status is probably related to a small fraction of tumor cells exhibiting MSI status (Table 5). Samples Ras-72 and Ras-78 were classified as MSS by ddPCR analysis but MSI-H by Promega analysis. When tested by Met-ddPCR, sample Ras-78 presented very low amount of cfDNA (0.5 ng/mL of plasma) with no detectable ctDNA, which is consistent with the MSS status by ddPCR analysis. Sample Ras-72 showed both high cfDNA and ctDNA load, while the historical IHC showed a pMMR determination. However, tissue sample was not available for retesting.

**Figure 4.**
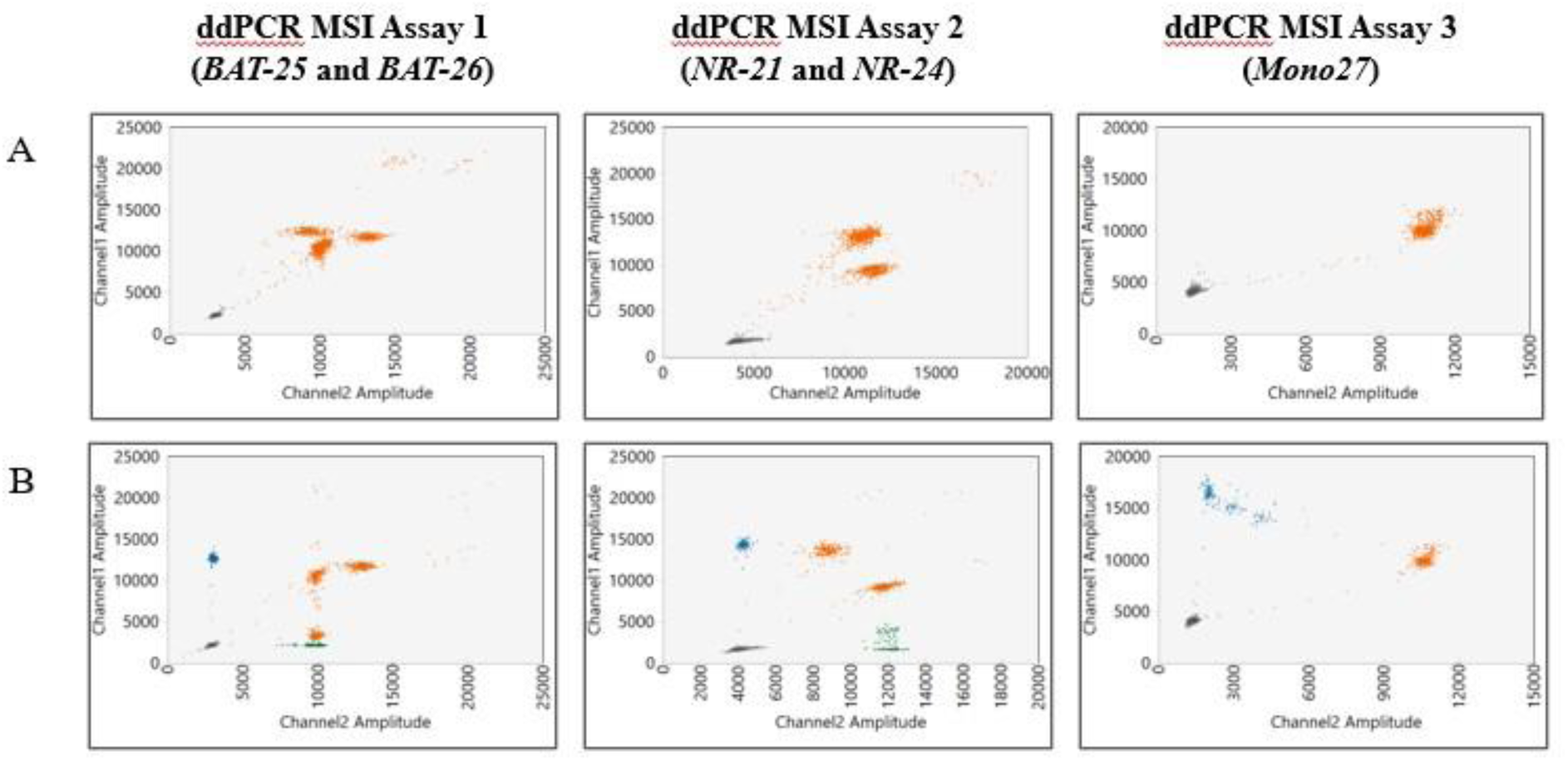
Example of ddPCR MSI 2-dimensional droplet plots of CRC cfDNA samples that are MSS (A, sample Ras-126) and MSI-H (B, sample Ras-21) samples.

**Table 4.**
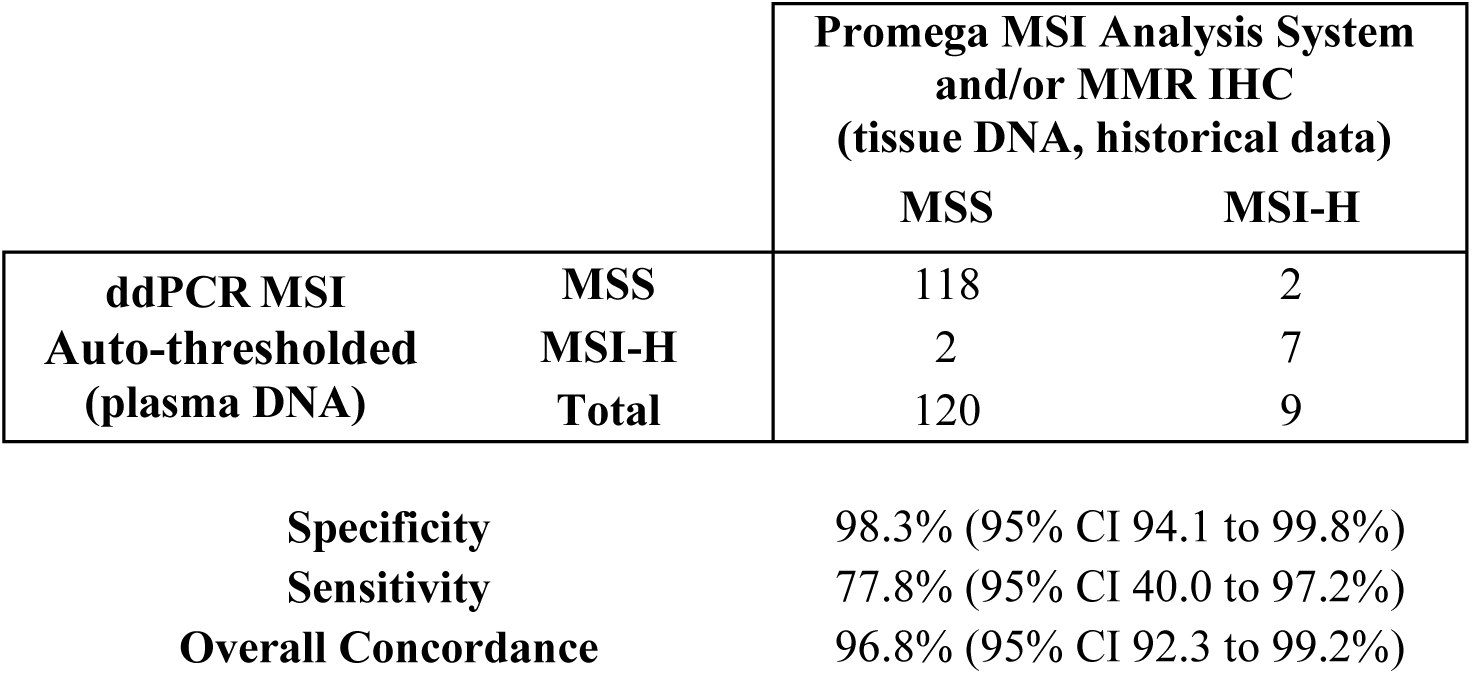
Concordance of cfDNA from RASANC cohort colorectal cancer patients between ddPCR MSI (cfDNA) and the Promega MSI Analysis System and/or IHC of matched tissue. The ddPCR MSI status results with manual thresholding were the same as with auto-thresholding. (MSS: microsatellite stable, MSI-H: microsatellite instability-high, MMR IHC: mismatch repair immunohistochemistry, ddPCR: droplet digital polymerase chain reaction.)

|  |  | **Promega MSI Analysis System**<br>**and/or MMR IHC**<br>**(tissue DNA, historical data)** |  |
| --- | --- | --- | --- |
|  |  | **MSS** | **MSI-H** |
| **ddPCR MSI** | **MSS** | 118 | 2 |
| **Auto-thresholded** | **MSI-H** | 2 | 7 |
| **(plasma DNA)** | **Total** | 120 | 9 |

|  |  |
| --- | --- |
| **Specificity** | 98.3% (95% CI 94.1 to 99.8%) |
| **Sensitivity** | 77.8% (95% CI 40.0 to 97.2%) |
| **Overall Concordance** | 96.8% (95% CI 92.3 to 99.2%) |

**Table 5.** Description of discordant cases in RASANC cohort. *In short, 1 mL of plasma was eluted from each patient in 50 uL of buffer solution and 18 uL tested for Met-ddCPR and 6.6 uL for MSI-ddPCR (see Material and Methods for details) (FF: Fresh Frozen, FFPE: formalin-fixed paraffin embedded, MSS: microsatellite stable, MSI-H: microsatellite instability-high, pMMR: proficient mismatch repair, dMMR: deficient mismatch repair, IHC: immunohistochemistry, cfDNA: circulating free DNA); NI: result that is not interpretable due to no ctDNA detection and/or too low cfDNA content.

| Sample | IHC MMR | Promega MSI Analysis system (tissue) | ddPCR MSI (cfDNA)* | Amplifiable cfDNA (Met-ddPCR)* | Presence of ctDNA (Met-ddPCR)* | Conclusion |
| --- | --- | --- | --- | --- | --- | --- |
| Ras-6 | pMMR | MSS | MSI-H (MSS on repeated test) | High (22.1 ng/mL) | Yes (115%) | Concordant data |
| Ras-39 | Not tested | MSS | MSI-H<br>Fractional Abundance (%):<br>*BAT-25* = 0.52<br>*BAT-26* = 0<br>*NR-21* = 0.20<br>*NR-24* = 0.20<br>*Mono-27* = 0.37 | High (21.5 ng/mL) | Yes (51.1%) | Discordant results may be due to tumor heterogeneity, where there's a small percent of tumor cells exhibiting MSI, as potentially indicated by the ddPCR MSI results. |
| Ras-72 | pMMR | MSI-H | MSS | High (36.2 ng/mL) | Yes (21.4%) | Concordant with IHC data only |
| Ras-78 | dMMR | MSI-H | MSS | Low (0.5 ng/mL) | No (low cfDNA) | NI |

## DISCUSSION

Microsatellite instability status determination is a crucial step for treatment management in cancer pathologies, including CRC. Association between MSI status and clinical outcomes has previously been described. In non-metastatic CRC, MSI-H tumors are associated with better prognosis than MSS tumors (20–22). Moreover, different responses to cancer treatments have also been observed depending on MSI status (23,24). Strong responses to immune checkpoint inhibitors have been observed for MSI-H CRC (6,25). In addition, MSI-H status is mandatory to identify Lynch Syndrome cases. In routine practice, MSI/MMR status is determined from tumor tissues using IHC or molecular testing. These methods present several limitations including low sensitivity (requiring 10 ng total DNA), high cost, long turn-around time and because they rely on localized tumor tissue analysis (requiring >20% tumor fraction) could lead to false negative results due to patient tumor heterogeneity or multiple tumor sites (26,27). Current methods are also limited in their scope to address MSI testing in plasma-based samples, being validated primarily for fresh frozen or FFPE tumor DNA.

MSI status testing is also becoming an available component of comprehensive genome profiling (CGP) or broad NGS panel testing from both technology product providers and clinical testing companies.

Although offering extensive multi-biomarker data simultaneously, CGP with NGS requires a higher bioinformatics burden and larger input DNA amounts (28). These requirements potentially preclude NGS based MSI testing from routine repeat testing for liquid biopsy analysis.

Recent years have seen large developments in minimally invasive liquid biopsy approaches especially for solid tumors (29). Such approaches could be of particular interest for the determination of MSI status in CRC tumors but most importantly for the patient monitoring during treatment and follow-up but require the development of highly sensitive techniques. To reach the required sensitivity, several approaches have been developed including mutant enrichment strategies, dedicated NGS approaches and more recently ddPCR strategies (11,20). Dedicated NGS approaches could lead to more broad, comprehensive results, for example by targeting multiple cancer types. However, when compared to ddPCR, NGS requires more complex data analysis, higher costs, longer turnaround time and has shown lower sensitivity (7,30).

Droplet-based digital PCR approaches have already demonstrated their ability to detect ctDNA in plasma with both high sensitivity and accuracy (13). Detection of tumor specific alterations in liquid biopsies by ddPCR has been largely demonstrated based on DNA variants (SNVs or indels), DNA methylation or copy number alterations. Very few studies have been reported on the development and/or validation of ddPCR approaches for MSI detection (11).

The present study describes the use and validation of a new Bio-Rad ddPCR MSI RUO Kit^®^ combined with an auto-calling algorithm based analysis software. Performance of the MSI ddPCR approach was compared to traditional MSI fragment analysis testing methods and IHC of MMR proteins on tumor.

Based on the duplicated analysis of DNA extracted from tumor tissues the developed ddPCR MSI approach demonstrated both high sensitivity of down to 1 ng of input DNA per reaction and efficiency of the auto-threshold calling. Among the 14 tested samples (FFPE and FF), 13 showed concordant MSI status at all DNA inputs when compared to historical determination (Promega and/or IHC). One sample presented discordant results at lower inputs likely due to low tumor content.

Validation of the assay performance was performed on tumor and plasma samples from 3 independent CRC patient cohorts. These samples consisted of 102 tumor tissues from the ALGECOLS multicentric cohort (stage I to III CRCs) and 129 plasma samples from the RASANC multicentric cohort (mCRCs) (9,16).

The technique in this work was first validated in DNA samples extracted from tumor tissue and demonstrated a high concordance level between MSI status determined by ddPCR analysis and historical data. The few discordant data were explained either by Promega MSI test misclassification or low tumor cell content within tested samples.

Initial validation of the ddPCR MSI kit with FFPE and FF samples of the ALGECOLS cohort shown a concordance of 92.1% with Promega MSI Analysis system, a sensitivity of 100% and an initial specificity of 91% when comparing with historical data. After discordant testing, only three discordant cases remained over 102 tested tissue samples leading to a specificity of 96.5% and a concordance of 97,1% between both methods. One of these discordances (case Alg-99) had 1 unstable marker in 5, indicating a borderline MSI status change that may have been ardently called on the initial Promega test. The other 2 discordant cases (Alg-25 and Alg-67) are likely due to low tumor content since fractional abundance for the 5 markers were very low (range: 1.14-1.72%). It is possible that these two borderline MSI-H samples by our ddPCR analysis, had been initially miss-classified MSS with the Promega MSI Analysis system because of its lower sensitivity. Indeed, this latter method requires a minimum of 20% tumor content to accurately determine MSI status. These results agree with the results from Gilson *et al.* that described, using 30 FFPE samples (15 CRC and 15 endometrial cancers), with the same 5 markers for ddPCR analysis, 100% overall agreement of ddPCR MSI analysis with Promega MSI system, with 88.4% sensitivity and 100% specificity. A similar result was observed by Bortolini Silveira *et al.* that analyzed 185 cancer samples, including 126 CRC tissues samples with a home-made drop-off MSI ddPCR assay (3-marker assay based on *BAT-26, ACVR2A* and *DEFB105A/B)* and obtained 100% sensitivity and specificity (31–33).

In the second part of the presented study the efficiency of the ddPCR MSI kit for the direct determination of MSI status within cfDNA was tested. Using 129 samples from the multicentric prospective RASANC cohort this work showed 96.9% concordant MSI determinations (125 samples). Among the 4 discordant cases, one sample (Ras-78) had very low levels of cfDNA, confirmed by Met-ddPCR, which was under the MSI ddPCR assay sensitivity level. For the 3 other samples (Ras-6, Ras-39, and Ras-72), cfDNA content was high according to Met-ddPCR. Ras-6 showed ∼50% fractional abundance for marker *BAT-25* at the first testing which was confirmed by a repeat ddPCR MSI test. However, for all other markers this sample showed droplet counts very close to the LOB, possibly indicating an MSS sample with a heterozygous locus for *BAT-25* only. The tumor of Ras-72 patient was called MSI-H by the Promega MSI system although its status was pMMR by IHC. The corresponding plasma sample was classified as MSS when tested by ddPCR MSI. This sample showed sufficient cfDNA and ctDNA levels for analysis. Finally, Ras-39 potentially illustrates the increased sensitivity of ddPCR MSI over Promega MSI Analysis system, with a total cfDNA input in the mid-range, and a relatively low fractional abundance for only 2/5 markers that still permitted auto-calling of an MSI-H status that would have been below the 20% fractional abundance required by Promega MSI Analysis system. The results presented here agree with the study of Bortolini Silveira *et al.*, that tested 72 plasma or serum samples (baseline samples and samples collected during treatments) corresponding to 14 MSI-H and 28 MSS tumor tissues. Interestingly, the plasma volume tested in our work was lower than in the Bortilini Silveira *et al*. study (33). This data confirmed the high sensitivity and accuracy of the developed assay even at low cfDNA content.

To confirm the interpretability of MSI status determination in plasma, WIF1/NPY Met-ddPCR was used to quantify both amplifiable cfDNA and ctDNA content within samples (9). The RASANC study, originally aimed to determine concordance between *RAS* mutation status in plasma and tumor of patients with mCRC. This methylation assay was used to determine the presence of ctDNA to ensure true negativity (i.e., wild-type *RAS* status) of tested samples. An accuracy of 94.8% for *RAS* mutation status was demonstrated, compared to 85.2% when no ctDNA quantification testing was performed (9). By confirming the presence and quantity of ctDNA within samples, the Met-ddPCR assay ensures true negativity, here MSS status.

The above sections illustrate potential limitations of the study including the potential for discordant (false positive) results determined by ddPCR compared to traditional methods due to the inherent higher sensitivity of the ddPCR technology. Comparison of results obtained from ddPCR amplification of FFPE derived DNA compared to plasma derived ctDNA may also be impacted by the differing nature of the two species of template, resulting in discordance, e.g., total DNA input, tumor DNA proportion, DNA fragment size and DNA integrity. Microsatellite instability in tumors with MSH6 deficiency specifically is not address by this study. Additionally, data presented is focused entirely on CRC samples and so further work would be required to show value for analysis of endometrial samples and other cancer types.

In conclusion, we developed and validated a highly sensitive ddPCR MSI assay and analysis. When tested on tumor tissues our methods showed high concordance with the Promega MSI system. The presented ddPCR analysis showed a higher sensitivity than the Promega MSI system for samples with low tumor content. The ability of the developed ddPCR MSI assay to efficiently determine MSI status in ctDNA extracted from plasma was also demonstrated. Combined with the developed auto-calling algorithm, this approach could find pertinent applications for diagnosis. Additional to its high performance, the developed ddPCR MSI assay has a shorter turnaround time compared to “standard” techniques. The ddPCR MSI assay may also be very useful for the analysis of plasma samples for patients with unavailable/interpretable tumor tissue analysis or for prospective analysis during treatment. Next step will be to evaluate patient monitoring with plasma ddPCR MSI as a predictor of treatment efficacy especially for MSI-H mCRC patients treated with immune checkpoint inhibitors.

## Availability of data

All data produced in the present study are available upon reasonable request to the authors

## Author contributions

K.C., C.E., J.A., S.G., A.D., D.LC., M.H., A.C., J.L., J.Y. performed the experiments and/or analyzed the data; D.T., P.L-P, V.T., M.H., A.C., H.B., S.G. designed the experiments and/or ensure supervision; D.T., P.L.P, V.T. secured fundings; A.Z., J-B.B., P.L-P, D.T., L.B., C.E. ensure patient accrual and data collection; C.E., V.T., L.B., P.L-P, D.T., M.H., A.C. ensure writing of the original draft; all authors contributed to the article and perform final approval of the article and its intellectual content. The authors also confirmed that questions related to the accuracy or integrity of the data and data analysis of the article are appropriately investigated.

## References

1. Arnold M, Sierra MS, Laversanne M, Soerjomataram I, Jemal A, Bray F: Global patterns and trends in colorectal cancer incidence and mortality. Gut 2017, 66(4):683–91

2. André T, Shiu KK, Kim TW, Jensen BV, Jensen LH, Punt C, Smith D, Garcia-Carbonero R, Benavides M, Gibbs P, de la Fouchardiere C, Rivera F, Elez E, Bendell J, Le DT, Yoshino T, Van Cutsem E, Yang P, Farooqui MZH, Marinello P, Diaz LA Jr: Pembrolizumab in Microsatellite-Instability–High Advanced Colorectal Cancer. N Engl J Med 2020, 383(23):2207–18

3. Hampel H, Frankel WL, Martin E, Arnold M, Khanduja K, Kuebler P, Clendenning M, Sotamaa K, Prior T, Westman JA, Panescu J, Fix D, Lockman J, LaJeunesse J, Comeras I, de la Chapelle A: Feasibility of Screening for Lynch Syndrome Among Patients With Colorectal Cancer. J Clin Oncol 2008, 26(35):5783–8

4. Coffin E, Dhooge M, Abou Ali E, Dermine S, Lavole J, Palmieri LJ, Chaussade S, Coriat R: Identification and management of patients with Lynch syndrome. Presse Med 2016, 48: 904–91

5. Guyot D’Asnières De Salins A, Tachon G, Cohen R, Karayan-Tapon L, Junca A, Frouin E, Godet J, Evrard C, Randrian V, Duval A, Svrcek M, Lascols O, Vignot S, Coulet F, André T, Fléjou JF, Cervera P, Tougeron D: Discordance between immunochemistry of mismatch repair proteins and molecular testing of microsatellite instability in colorectal cancer. ESMO Open 2021, 6(3):100120

6. Evrard C, Messina S, Sefrioui D, Frouin É, Auriault ML, Chautard R, Zaanan A, Jaffrelot M, De La Fouchardière C, Aparicio T, Coriat R, Godet J, Silvain C, Randrian V, Sabourin JC, Guimbaud R, Miquelestorena-Standley E, Lecomte T, Moulin V, Karayan-Tapon L, Tachon G, Tougeron D: Heterogeneity of Mismatch Repair Status and Microsatellite Instability between Primary Tumour and Metastasis and Its Implications for Immunotherapy in Colorectal Cancers. Int J Mol Sci 2022, 23(8):4427

7. Nadal C, Winder T, Gerger A, Tougeron D: Future perspectives of circulating tumor DNA in colorectal cancer. Tumour Biol 2017, 39(5):1010428317705749

8. Anker P, Lefort F, Vasioukhin V, Lyautey J, Lederrey C, Chen XQ, Stroun M, Mulcahy HE, Farthing MJ: K-ras mutations are found in DNA extracted from the plasma of patients with colorectal cancer. Gastroenterology 1997, 112(4):1114–20

9. Bachet JB, Bouché O, Taieb J, Dubreuil O, Garcia ML, Meurisse A, Normand C, Gornet JM, Artru P, Louafi S, Bonnetain F, Thirot-Bidault A, Baumgaertner I, Coriat R, Tougeron D, Lecomte T, Mary F, Aparicio T, Marthey L, Taly V, Blons H, Vernerey D, Laurent-Puig P: RAS mutation analysis in circulating tumor DNA from patients with metastatic colorectal cancer: the AGEO RASANC prospective multicenter study. Ann Oncol 2018, 29(5):1211–9

10. Nakamura Y, Okamoto W, Denda T, Nishina T, Komatsu Y, Yuki S, Yasui H, Esaki T, Sunakawa Y, Ueno M, Shinozaki E, Matsuhashi N, Ohta T, Kato K, Ohtsubo K, Bando H, Hara H, Satoh T, Yamazaki K, Yamamoto Y, Okano N, Terazawa T, Kato T, Oki E, Tsuji A, Horita Y, Hamamoto Y, Kawazoe A, Nakajima H, Nomura S, Mitani R, Yuasa M, Akagi K, Yoshino T: Clinical Validity of Plasma-Based Genotyping for Microsatellite Instability Assessment in Advanced GI Cancers: SCRUM-Japan GOZILA Substudy. JCO Precis Oncol 2022, 6:e2100383

11. Silveira AB, Bidard FC, Kasperek A, Melaabi S, Tanguy ML, Rodrigues M, Bataillon G, Cabel L, Buecher B, Pierga JY, Proudhon C, Stern MH: High-Accuracy Determination of Microsatellite Instability Compatible with Liquid Biopsies. Clin Chem 2020, 66(4):606–613

12. Hudecova I: Digital PCR analysis of circulating nucleic acids. Clin Biochem 2015, 48(15):948–56

13. Postel M, Roosen A, Laurent-Puig P, Taly V, Wang-Renault SF: Droplet-based digital PCR and next generation sequencing for monitoring circulating tumor DNA: a cancer diagnostic perspective. Expert Rev Mol Diagn 2018, 18(1):7–17

14. Pekin D, Skhiri Y, Baret JC, Le Corre D, Mazutis L, Salem CB, Millot F, El Harrak A, Hutchison JB, Larson JW, Link DR, Laurent-Puig P, Griffiths AD, Taly V: Quantitative and sensitive detection of rare mutations using droplet-based microfluidics. Lab Chip 2011, 11(13):2156–66

15. Boland CR, Thibodeau SN, Hamilton SR, Sidransky D, Eshleman JR, Burt RW, Meltzer SJ, Rodriguez-Bigas MA, Fodde R, Ranzani GN, Srivastava S: A National Cancer Institute Workshop on Microsatellite Instability for Cancer Detection and Familial Predisposition: Development of International Criteria for the Determination of Microsatellite Instability in Colorectal Cancer. Cancer Res 1998, 58(22):5248–57

16. Benhaim L, Bouché O, Normand C, Didelot A, Mulot C, Le Corre D, Garrigou S, Djadi-Prat J, Wang-Renault SF, Perez-Toralla K, Pekin D, Poulet G, Landi B, Taieb J, Selvy M, Emile JF, Lecomte T, Blons H, Chatellier G, Link DR, Taly V, Laurent-Puig P: Circulating tumor DNA is a prognostic marker of tumor recurrence in stage II and III colorectal cancer: multicentric, prospective cohort study (ALGECOLS). Eur J Cancer 2021, 159:24–33

17. dMIQE Group; Huggett JF: The Digital MIQE Guidelines Update: Minimum Information for Publication of Quantitative Digital PCR Experiments for 2020. Clin Chem 2020, 66(8):1012–1029

18. Garlan F, Laurent-Puig P, Sefrioui D, Siauve N, Didelot A, Sarafan-Vasseur N, Michel P, Perkins G, Mulot C, Blons H, Taieb J, Di Fiore F, Taly V, Zaanan A: Early Evaluation of Circulating Tumor DNA as Marker of Therapeutic Efficacy in Metastatic Colorectal Cancer Patients (PLACOL Study). Clin Cancer Res 2017, 23(18):5416–5425

19. Taieb J, Taly V, Henriques J, Bourreau C, Mineur L, Bennouna J, Desrame J, Louvet C, Lepere C, Mabro M, Egreteau J, Bouche O, Mulot C, Hormigos K, Chaba K, Mazard T, de Gramont A, Vernerey D, André T, Laurent-Puig P: Prognostic Value and Relation with Adjuvant Treatment Duration of ctDNA in Stage III Colon Cancer: a Post Hoc Analysis of the PRODIGE-GERCOR IDEA-France Trial. Clin Cancer Res 2021, 27(20):5638–5646

20. Yu F, Makrigiorgos A, Leong KW, Makrigiorgos GM: Sensitive detection of microsatellite instability in tissues and liquid biopsies: Recent developments and updates. Comput Struct Biotechnol J 2021, 19:4931–4940

21. Tougeron D, Sickersen G, Mouillet G, Zaanan A, Trouilloud I, Coriat R, Aparicio T, Des Guetz G, Lecaille C, Artru P, Cauchin E, Sefrioui D, Boussaha T, Ferru A, Matysiak-Budnik T, Silvain C, Karayan-Tapon L, Pagès JC, Vernerey D, Bonnetain F, Michel P, Taïeb J, Lecomte T; Association des Gastro-Entérologues Oncologues (AGEO): Predictors of disease-free survival in colorectal cancer with microsatellite instability: An AGEO multicentre study. Eur J Cancer 2015, 51(8):925–34

22. Gallois C, Laurent-Puig P, Taieb J: Methylator phenotype in colorectal cancer: A prognostic factor or not? Crit Rev Oncol Hematol 2016, 99:74–80

23. Vilar E, Gruber SB: Microsatellite instability in colorectal cancer—the stable evidence. Nat Rev Clin Oncol 2010, 7(3):153–62

24. Tougeron D, Sueur B, Zaanan A, de la Fouchardiére C, Sefrioui D, Lecomte T, Aparicio T, Des Guetz G, Artru P, Hautefeuille V, Coriat R, Moulin V, Locher C, Touchefeu Y, Lecaille C, Goujon G, Ferru A, Evrard C, Chautard R, Gentilhomme L, Vernerey D, Taieb J, André T, Henriques J, Cohen R; Association des Gastro-entérologues Oncologues (AGEO): Prognosis and chemosensitivity of deficient MMR phenotype in patients with metastatic colorectal cancer: An AGEO retrospective multicenter study. Int J Cancer 2020, 147(1):285–296

25. André T, Shiu KK, Kim TW, Jensen BV, Jensen LH, Punt C, Smith D, Garcia-Carbonero R, Benavides M, Gibbs P, de la Fouchardiere C, Rivera F, Elez E, Bendell J, Le DT, Yoshino T, Van Cutsem E, Yang P, Farooqui MZH, Marinello P, Diaz LA Jr; KEYNOTE-177 Investigators: Pembrolizumab in Microsatellite-Instability–High Advanced Colorectal Cancer. N. Engl. J. Med 2020, 383: 2207–2218

26. Wang Y, Shi C, Eisenberg R, Vnencak-Jones CL: Differences in Microsatellite Instability Profiles between Endometrioid and Colorectal Cancers. J Mol Diagn 2017, 19(1):57–64

27. Tachon G, Frouin E, Karayan-Tapon L, Auriault ML, Godet J, Moulin V, Wang Q, Tougeron D: Heterogeneity of mismatch repair defect in colorectal cancer and its implications in clinical practice. Eur J Cancer 2018, 95:112–116

28. Wang Z, Zhao X, Gao C, Gong J, Wang X, Gao J, Li Z, Wang J, Yang B, Wang L, Zhang B, Zhou Y, Wang D, Li X, Bai Y, Li J and Shen L. Plasma-based microsatellite instability detection strategy to guide immune checkpoint blockade treatment. J Immunother Cancer 2020,8(2):e001297

29. Poulet G, Massias J, Taly V: Liquid Biopsy: General Concepts. Acta Cytol 2019, 63(6):449–455

30. Willis J, Lefterova MI, Artyomenko A, Kasi PM, Nakamura Y, Mody K, Catenacci DVT, Fakih M, Barbacioru C, Zhao J, Sikora M, Fairclough SR, Lee H, Kim KM, Kim ST, Kim J, Gavino D, Benavides M, Peled N, Nguyen T, Cusnir M, Eskander RN, Azzi G, Yoshino T, Banks KC, Raymond VM, Lanman RB, Chudova DI, Talasaz A, Kopetz S, Lee J, Odegaard JI: Validation of Microsatellite Instability Detection. Using a Comprehensive Plasma-Based Genotyping Panel. Clin Cancer Res 2019, 25(23):7035–7045

31. Gilson P, Levy J, Rouyer M, Demange J, Husson M, Bonnet C, Salleron J, Leroux A, Merlin JL, Harlé A: Evaluation of 3 molecular-based assays for microsatellite instability detection in formalin-fixed tissues of patients with endometrial and colorectal cancers. Sci Rep 2020, 10(1):16386

32. Long DR, Waalkes A, Panicker VP, Hause RJ, Salipante SJ: Identifying Optimal Loci for the Molecular Diagnosis of Microsatellite Instability. Clin Chem 2020, 66(10):1310–1318

33. Bortolini Silveira A, Bidard FC, Tanguy ML, Girard E, Trédan O, Dubot C, et al. Multimodal liquid biopsy for early monitoring and outcome prediction of chemotherapy in metastatic breast cancer. NPJ Breast Cancer (2021)7(1):115.doi: 10.1038/s41523-021-00319-4.

