## Supplemental data for "Detection of Microsatellite Instability in Colorectal Cancer Tissue and Plasma samples using a new Multiplex Droplet Digital PCR kit"

**New Multiplex Droplet Digital PCR for highly sensitive detection of microsatellite instability**

### Supplementary Data

**Supplementary Table 1. Digital MIQE2020 guidelines (dMIQE2020) checklist.**

| ITEM TO CHECK | PROVIDED | COMMENT |
|---|---|---|
| | Y/N | |
| **1. SPECIMEN** | | |
| Detailed description of specimen type and numbers | Y | Materials and Methods, references |
| Sampling procedure (including time to storage) | Y | Materials and Methods, references |
| Sample aliquotation, storage conditions and duration | Y | Materials and Methods, references |
| **2. NUCLEIC ACID EXTRACTION** | | |
| Description of extraction method including amount of sample processed | Y | Materials and Methods, references |
| Volume of solvent used to elute/resuspend extract | Y | Materials and Methods, references |
| Number of extraction replicates | Y | Materials and Methods, references |
| Extraction blanks included? | N | |
| **3. NUCLEIC ACID ASSESSMENT AND STORAGE** | | |
| Method to evaluate quality of nucleic acids | Y | Materials and Methods |
| Method to evaluate quantity of nucleic acid | Y | Materials and Methods |
| Storage conditions: temperature, concentration, duration, buffer, aliquots | N | |
| Clear description of dilution steps used to prepare working DNA solution | N | |
| **4. NUCLEIC ACID MODIFICATION** | | |

**New Multiplex Droplet Digital PCR for highly sensitive detection of microsatellite instability**

| | | |
|---|---|---|
| Template modification (digestion, sonication, pre-amplification, bisulfite etc.) | Y | Bisulfite conversion: Materials and Methods, references |
| Re-extraction performed? | Y | Materials and Methods, references |
| **5. REVERSE TRANSCRIPTION** | N | |
| cDNA priming method + concentration | | Not Applicable |
| One or two step protocol (include reaction details for two step) | | Not Applicable |
| Amount of RNA used per reaction | | Not Applicable |
| Detailed reaction components and conditions | | Not Applicable |
| Estimated copies measured with and without addition of RT | | Not Applicable |
| Manufacturer of reagents used and catalogue number | | Not Applicable |
| Storage of cDNA: temperature, concentration, duration, buffer, aliquots | | Not Applicable |
| **6. dPCR OLIGONUCLEOTIDES AND TARGET INFORMATION** | | |
| Sequence accession number | N | Commercial kit: Bio-Rad ddPCR MSI RUO Kit |
| Location of amplicon | N | Commercial kit: Bio-Rad ddPCR MSI RUO Kit |
| Amplicon length | N | Commercial kit: Bio-Rad ddPCR MSI RUO Kit |
| Primer and probe sequences (or amplicon context sequence) | N | BAT-25<br><br>TAAATTAGAACAGGAATTCCAAAGAGACAGCAGTT<br>GGAACATGAAGAAACTAAATTTCCTCTGCTTTTGGT<br>TACCACACTTCAAAATGACATTCTGCATTTTAACTA<br>TGGCTCTAAAATGCTCTGTTCTCAAAAAAAAAAAA<br>AAAAATCAAAAAAACAAAACACAAAACTCTTTAGA<br>GAATCACTCCCACTTACATTCTTGGAGGCGAGGAA<br>AGCCATGCCCTTTGCCACCTGGTAAGAAAAGCTCA<br>GCAAGTCTTCTAAGTCTAGGGCCAACTCGTCATCCT<br>CCATGATGGCGGGAGTCACATCTCTTTCTAT<br><br>BAT-26 |

| | | |
|---|---|---|
| | | TTAGAACTCTTATCAGATGATTCCAACTTTGGACAG<br>TTTGAACTGACTACTTTTGACTTCAGCCAGTATATG<br>AAATTGGATATTGCAGCAGTCAGAGCCCTTAACCT<br>TTTTCAGGTAAAAAAAAAAAAAAAAAAAAGGGTTAA<br>AAATGTTGAATGGTTAAAAAATGTTTTCATTGACAT<br>ATACTGAAGAAGCTTATAAAGGAGCTAAAATATTT<br>TGAAATATTATTATACTTGGATTAGATAACTAGCTT<br>TAAATGGCTGTATTTTTCTCTCCCCTCCTCCACTCCA<br>CTTTTTAACTTTT<br><br>NR-21<br><br>TGCTACTCTCTAAAAAAGGCAAGCAGATAAAAGAG<br>AACACGAAAAATATTCCTACTCCGCATTCACACTTT<br>CTGGTCACTCGCGTTTACAAACAAGAAAAGTGTTG<br>CTAAAAAAAAAAAAAAGGCCAGGGGAGACATACAT<br>TTAAATATAAAAATAGAACTGTGCCAGCGACTCCG<br>GCTGGAATTCTGCTGAAAGGGATGTGTCTTCAGAA<br>ACCAGGATGGTTCTG<br><br>NR-24<br><br>GGCTGAGGCAGGAGAATGGCGTGAACCCGGGAGG<br>CGGACGTTGCAGTGAGCGGAGATTGTGCCATTGCA<br>TTCCAACCTGGGTGACAGAGTGAGACTCTGTCTCA<br>CAAAAAAAAAAAAAAAAAATAGGACTGGGCCCAGGG<br>AAGAGAAGAGTTTTTGGAGTCAGGAGGTAAAATTC<br>AGCAATGGGGATCACGTTAAGGCAGACCTTAAATA<br>TTCAGATAATTTTTCACTAAATAGGGAGCTATTTTA<br>GGTCTTGAGGAAGGGCAGTGATGGGCAGGAAGGG<br>GTCATTTG<br><br>Mono-27:<br>GGCAGGGAAATGGTGGGAACCCAGGGGGTGGAGA<br>TTGCAGTGAGCTGAGATTGCGCCACTGCACTCCAG<br>CGTGGGAGACAGAGCAAGACTCTGCCTCAAAAAAA<br>AAAAAAAAAAAAATCCTGGTTTTACTTTTTTTTCTTT<br>TTTAGTTGGCCAAGTGAAATTTGATCCACCCTTAAG<br>AAAGGAGACAGAACCACATCATGAACTTGTAAGTA<br>GTATAGCCTTAGAATGTTAGCACTGAAAATAAATG<br>TTTTAATTTGTTTTTGTGTGATGTATTAGTACTGACC<br>AAAAAAGCT |
| Location and identity of any modifications | N | Commercial kit: Bio-Rad ddPCR MSI RUO Kit |
| Manufacturer of oligonucleotides | N | Commercial kit: Bio-Rad ddPCR MSI RUO Kit |
| **7. dPCR PROTOCOL** | | |
| Manufacturer of dPCR instrument and instrument model | Y | Bio-Rad Laboratories QXDx ddPCR System |

**New Multiplex Droplet Digital PCR for highly sensitive detection of microsatellite instability**

| | | |
|---|---|---|
| Buffer/kit Catalogue No and manufacturer | Y | Commercial kit: Bio-Rad ddPCR MSI RUO Kit |
| Primer and probe concentration | N | |
| Pre-reaction volume and composition (incl. proportion of template added) | Y | 6.6ul sample per 22ul reaction |
| Template treatment (initial heating or chemical denaturation) | N | |
| Polymerase identity and concentration, Mg++ and dNTP concentrations | N | |
| Complete thermocycling parameters | Y | Materials and Methods |
| **8. ASSAY VALIDATION** | | |
| Details of optimisation performed | | Bio-Rad ddPCR MSI RUO Kit User Guide |
| Analytical specificity (vs. related sequences) | | Bio-Rad ddPCR MSI RUO Kit User Guide |
| Analytical sensitivity (LoD) | Y | Bio-Rad ddPCR MSI RUO Kit User Guide |
| Testing for inhibitors (from biological matrix/extraction) | N | Not performed |
| **9. DATA ANALYSIS** | | |
| Description of dPCR experimental design | Y | Materials and Methods |
| Comprehensive details +ve and -ve of controls (whether applied for QC or for estimation of error) | Y | Materials and Methods: positive and negative controls included as part of kit on every plate. |
| Examples of +ve and -ve experimental results (in supplemental materials) | Y | In results |
| Description of technical replication | Y | Materials and Methods |
| Repeatability (intra-experiment variation) | Y | Materials and Methods |
| Reproducibility (inter-experiment/user/lab etc. variation) | N | |

**New Multiplex Droplet Digital PCR for highly sensitive detection of microsatellite instability**

| | | |
|---|---|---|
| Number of partitions measured (average and standard deviation) | Y | Materials and Methods |
| Partition volume used | N | |
| Copies per partition (λ or equivalent) (average and standard deviation) | N | |
| dPCR analysis program (source, version) | Y | QuantaSoft Analysis Pro v1.0 (manual threshold)<br>QX Manager Premium Edition v2.0.0.456 (auto-threshold) |
| Description of normalisation method | N | Material and Methods: dPCR manual thresholding and auto-thresholding was normalized to the mean cluster densities of the positive controls run on every plate. |
| Statistical methods used for analysis | N | A Poisson distribution analysis was performed on the dPCR data to calculate copies per ul. |
| Data submission using RDML | N | Availability of data upon request following all necessary regulatory requirements. |

**New Multiplex Droplet Digital PCR for highly sensitive detection of microsatellite instability**

**Supplementary Table 2. Validation results obtained for the analysis of FFPE samples using the auto-thresholding algorithm.** Of the over 200 contrived and CRC patient FF, FFPE and cfDNA samples used to develop the auto-thresholding algorithm 60 were FFPE (47 MSI-H and 13 MSS) and 22 were cfDNA (14 MSI-H and 8 MSS), and were used to train the neural network. Another 94 FFPE, 24 MSI-H and 70 MSS, were used to validate the auto-thresholding algorithm. The validation results are shown in the table.

| Sample Type | n | Vendor MSI Status* | Concordant Samples by ddPCR MSI | Sensitivity or Specificity | Overall Concordance |
|---|---|---|---|---|---|
| FFPE | 24 | MSI-H | 23 | 95.8% | 96.4% |
| | 70** | MSS | 67 | 95.7% | |

*Orthogonal data is immunohistochemistry of mismatch repair proteins

**20 are FFPE of colon tissue from normal patients and presumed MSS

**New Multiplex Droplet Digital PCR for highly sensitive detection of microsatellite instability**

**Supplementary Table 3.** Summary of all ddPCR Microsatellite Instability (MSI) RUO kit data that was excluded from the analysis due to a failure or low droplet count of <10,000.

| Sample | Sample Type | ddPCR MSI Assay | Reason for Exclusion |
|---|---|---|---|
| Alg-10 | FF/FFPE | 1 | Low droplet count of 4698 |
| Alg-11 | FF/FFPE | 2 | Low droplet count of 8603 |
| Alg-14 | FF/FFPE | 3 | Low droplet count of 2006 |
| Alg-14 | FF/FFPE | 2 | Unsuccessful droplet generation, no droplets detected |
| Alg-16 | FF/FFPE | 1 | Low droplet count of 1396 |
| Alg-16 | FF/FFPE | 2 | Unsuccessful droplet generation |
| Alg-23 | FF/FFPE | 1 | Low droplet count of 3699 |
| Alg-28 | FF/FFPE | 2 | Unsuccessful droplet generation, no droplets detected |
| Alg-34 | FF/FFPE | 3 | Low droplet count of 3342 |
| Alg-51 | FF/FFPE | 2 | Unsuccessful droplet generation |
| Alg-58 | FF/FFPE | 2 | Low droplet count of 3047 |
| Alg-58 | FF/FFPE | 3 | Low droplet count of 5056 |
| Alg-61 | FF/FFPE | 3 | Low droplet count of 2816 |
| Alg-67 | FF/FFPE | 2 | Low droplet count of 5493 |
| Alg-70 | FF/FFPE | 3 | Low droplet count of 5938 |
| Alg-73 | FF/FFPE | 1 | Insufficient plate seal before thermal cycling |
| Alg-91 | FF/FFPE | 3 | Low droplet count of 7958 |
| Alg-93 | FF/FFPE | 1 | Low droplet count of 675 |
| Ras-95 | cfDNA | 3 | Cluster mirroring |

**New Multiplex Droplet Digital PCR for highly sensitive detection of microsatellite instability**

**Supplementary Table 4.** Concordance of FF and FFPE tissues from colorectal cancer patients between auto-thresholded ddPCR MSI with the Promega MSI Analysis System v1.2 (**A**). There were twelve samples that were not concordant for all markers: Samples [†]Alg-96; [*]Alg-25, Alg-67; [ǂ]Alg-99, [‖]Alg-45, [¶]Alg-30, Alg-43, Alg-53, Alg-63; [**]Alg-17; [††]Alg-88; [§]Alg-28. For Alg-28, the same 4 markers were not unstable for both ddPCR MSI and the MSI Analysis System v1.2. Eleven samples were re-tested with the MSI Analysis System v1.2 (Alg-67 not re-tested) (**B**). Seven of these were concordant compared to ddPCR MSI, while four remained discordant by marker: Sample [*]Alg-25 & Alg-67, [ǂ]Alg-99, [¶]Alg-30, [§]Alg-28.

**A**

| | | | **Promega MSI Analysis (tissue DNA), Historical** | | | | | |
|---|---|---|---|---|---|---|---|---|
| | | | **MSS** | | **MSI-H** | | | |
| | | | 0 | 1 | 2 | 3 | 4 | 5 |
| **ddPCR MSI Auto-threshold (tissue DNA)** | **MSS** | 0 | 83 | | | | | |
| | | 1 | 1[†] | | | | | |
| | **MSI-H** | 2 | 2[*] | | | | | |
| | | 3 | 1[ǂ] | | | | | |
| | | 4 | | | | | 1[§] | |
| | | 5 | 1[‖] | 4[¶] | 1[*] | | 1[†] | 7 |

**B**

| | | | **Promega MSI Analysis + Discordant Repeat Result (tissue DNA), Historical** | | | | | |
|---|---|---|---|---|---|---|---|---|
| | | | **MSS** | | **MSI-H** | | | |
| | | | 0 | 1 | 2 | 3 | 4 | 5 |
| **ddPCR MSI Auto-threshold (tissue DNA)** | **MSS** | 0 | 83 | | | | | |
| | | 1 | | 1 | | | | |
| | **MSI-H** | 2 | 2[*] | 1[ǂ] | | | | |
| | | 3 | | | | | | |
| | | 4 | | | | | | 1[§] |
| | | 5 | | | | 1[¶] | | 13 |

**New Multiplex Droplet Digital PCR for highly sensitive detection of microsatellite instability**

**Supplementary Table 5**. Concordance of FF and FFPE tissues from colorectal cancer patients between manual thresholded ddPCR MSI with the Promega MSI Analysis System v1.2 (**A**). There were fourteen samples that were not concordant for all markers: Sample [†]Alg-1, Alg-67, Alg-96, Alg-99; [*]Alg25; [ǀ]Alg-45; [¶]Alg-30, Alg-43; [**]Alg-53, Alg-63; [ǂ]Alg-17; [§]Alg-28; [††]Alg-88; [#]Alg-75. For Sample Alg-28, the same 4 markers were not unstable for both ddPCR MSI and the MSI Analysis System v1.2. Twelve of the fourteen samples were re-tested with the MSI Analysis System v1.2 (**B**). Sample Alg-1 and Alg-67 not retested. Seven of these were concordant compared to ddPCR MSI, while five remained discordant by marker: Sample [†]Alg-1, Alg-67; *Alg-25; [¶]Alg-30, [ǂ]Alg-75; [§]Alg-28, Alg-43. The concordance by MSI status was high at 101 of 102 (**C**).

**A**

**Promega MSI Analysis (tissue DNA), Historical**

| | | | **MSS** | | **MSI-H** | | | |
|---|---|---|---|---|---|---|---|---|
| | | | 0 | 1 | 2 | 3 | 4 | 5 |
| **ddPCR MSI Manual Threshold (tissue DNA)** | **MSS** | 0 | 82 | | | | | |
| | | 1 | 4[†] | | | | | |
| | **MSI-H** | 2 | 1[*] | | | | | |
| | | 3 | | | | | | 1[#] |
| | | 4 | | 2[¶] | | | 1[§] | |
| | | 5 | 1[ǀ] | 2[*] | 1[ǂ] | | 1[†] | 6 |

**B**

**Promega MSI Analysis + Discordant Repeat Result (tissue DNA), Historical**

| | | | **MSS** | | **MSI-H** | | | |
|---|---|---|---|---|---|---|---|---|
| | | | 0 | 1 | 2 | 3 | 4 | 5 |
| **ddPCR MSI Manual Threshold (tissue DNA)** | **MSS** | 0 | 82 | | | | | |
| | | 1 | 2[†] | 2 | | | | |
| | **MSI-H** | 2 | 1[*] | | | | | |
| | | 3 | | | | | | 1[ǂ] |
| | | 4 | | | | 1[¶] | | 2[§] |
| | | 5 | | | | | | 11 |

**New Multiplex Droplet Digital PCR for highly sensitive detection of microsatellite instability**

**C**

| | | Promega MSI Analysis system + Discordant Repeat Test | |
|---|---|---|---|
| | | **MSS** | **MSI-H** |
| **ddPCR MSI Manual Threshold** | **MSS** | 86 | 0 |
| | **MSI-H** | 1 | 15 |

**New Multiplex Droplet Digital PCR for highly sensitive detection of microsatellite instability**

**Supplementary Table 6**. Comparison of ddPCR MSI with auto-thresholding for 14 FF and FFPE samples when tested at an input per well of 18, 5 and 1 ng/well, measured by Qubit dsDNA BR Assay. Seven (7) of the samples were determined as MSS and 7 as MSI-H by the Promega MSI Analysis System. The ddPCR MSI results reflect data for replicate 2 of a total of 2 replicates.

| | Promega MSI Analysis | | | | | | ddPCR MSI - 18 ng/rxn | | | | | | ddPCR MSI - 5 ng/rxn | | | | | | ddPCR MSI - 1 ng/rxn | | | | | |
|---|---|---|---|---|---|---|---|---|---|---|---|---|---|---|---|---|---|---|---|---|---|---|---|---|
| Sample | BAT-25 | BAT-26 | NR-21 | NR-24 | Mono-27 | Status | BAT-25 | BAT-26 | NR-21 | NR-24 | Mono-27 | Status | BAT-25 | BAT-26 | NR-21 | NR-24 | Mono-27 | Status | BAT-25 | BAT-26 | NR-21 | NR-24 | Mono-27 | Status |
| Alg-1 | - | - | - | - | - | MSS | - | - | - | - | - | MSS | - | - | - | + | - | MSS | - | - | - | - | - | MSS |
| Alg-2 | - | - | - | - | - | MSS | - | - | - | - | - | MSS | - | - | - | - | - | MSS | - | - | - | - | + | MSS |
| Alg-14 | - | - | - | - | - | MSS | - | - | na | na | na | na | - | - | - | - | - | MSS | - | - | - | - | - | MSS |
| Alg-16 | - | - | - | - | - | MSS | na | na | na | na | - | na | - | - | - | - | - | MSS | - | - | - | - | - | MSS |
| Alg-67 | - | - | - | - | - | MSS | - | + | na | na | - | na | - | + | - | - | - | MSS | - | - | - | - | + | MSS |
| Alg-87 | - | - | - | - | - | MSS | - | - | - | - | - | MSS | - | - | - | - | - | MSS | - | - | - | - | - | MSS |
| Alg-100 | - | - | - | - | - | MSS | - | - | - | - | - | MSS | - | - | - | - | - | MSS | - | - | - | - | - | MSS |
| Alg-28 | + | + | + | + | + | MSI-H | + | + | + | + | - | MSI-H | + | + | + | + | - | MSI-H | + | + | + | + | + | MSI-H |
| Alg-30 | + | + | + | - | - | MSI-H | + | + | + | + | + | MSI-H | + | + | + | + | + | MSI-H | + | + | + | + | + | MSI-H |
| Alg-33 | + | + | + | + | + | MSI-H | + | + | + | + | + | MSI-H | + | + | + | + | + | MSI-H | + | + | + | + | + | MSI-H |
| Alg-43 | + | + | + | + | + | MSI-H | + | + | + | + | + | MSI-H | + | + | + | + | + | MSI-H | + | + | + | + | + | MSI-H |
| Alg-58 | + | + | + | + | + | MSI-H | + | + | na | na | na | na | + | + | + | + | + | MSI-H | + | + | + | + | + | MSI-H |
| Alg-83 | + | + | + | + | + | MSI-H | + | + | + | + | + | MSI-H | + | + | + | + | + | MSI-H | + | + | + | + | + | MSI-H |
| Alg-89 | + | + | + | + | + | MSI-H | + | + | + | + | + | MSI-H | + | + | + | + | + | MSI-H | + | + | + | + | + | MSI-H |

| | |
|---|---|
| - | stable |
| + | unstable |
| na | failed |

**New Multiplex Droplet Digital PCR for highly sensitive detection of microsatellite instability**

**Supplementary Table 7.** Comparison of ddPCR MSI with manual thresholding for 14 FF and FFPE samples when tested at an input per well of 18, 5 and 1 ng/well, measured by Qubit dsDNA BR Assay. Seven (7) of the samples were determined as MSS and 7 as MSI-H by the Promega MSI Analysis System. The ddPCR MSI results reflect data for replicate 1 of a total of 2 replicates.

| | Promega MSI Analysis | | | | | | ddPCR MSI - 18 ng/rxn | | | | | | ddPCR MSI - 5 ng/rxn | | | | | | ddPCR MSI - 1 ng/rxn | | | | | |
|---|---|---|---|---|---|---|---|---|---|---|---|---|---|---|---|---|---|---|---|---|---|---|---|---|
| Sample | BAT-25 | BAT-26 | NR-21 | NR-24 | Mono-27 | Status | BAT-25 | BAT-26 | NR-21 | NR-24 | Mono-27 | Status | BAT-25 | BAT-26 | NR-21 | NR-24 | Mono-27 | Status | BAT-25 | BAT-26 | NR-21 | NR-24 | Mono-27 | Status |
| Alg-1 | - | - | - | - | - | MSS | - | + | - | - | - | MSS | - | - | - | - | - | MSS | - | - | - | - | - | MSS |
| Alg-2 | - | - | - | - | - | MSS | - | - | - | - | - | MSS | - | - | - | - | - | MSS | - | - | - | - | - | MSS |
| Alg-14 | - | - | - | - | - | MSS | - | - | - | - | - | MSS | - | - | - | - | - | MSS | - | - | - | - | - | MSS |
| Alg-16 | - | - | - | - | - | MSS | - | - | - | - | - | MSS | - | - | - | - | - | MSS | - | - | - | - | - | MSS |
| Alg-67 | - | - | - | - | - | MSS | - | + | - | - | - | MSI-H | - | + | - | - | - | MSS | - | - | - | - | - | MSS |
| Alg-87 | - | - | - | - | - | MSS | - | - | - | - | - | MSS | - | - | - | - | - | MSS | - | - | - | - | - | MSS |
| Alg-100 | - | - | - | - | - | MSS | - | - | - | - | - | MSS | - | - | - | - | - | MSS | - | - | - | - | - | MSS |
| Alg-28 | + | + | + | + | + | MSI-H | + | + | + | + | - | MSI-H | + | + | + | + | - | MSI-H | - | + | + | + | - | MSI-H |
| Alg-30 | + | + | + | - | - | MSI-H | + | + | + | - | + | MSI-H | + | + | - | - | + | MSI-H | + | + | + | + | - | MSI-H |
| Alg-33 | + | + | + | + | + | MSI-H | + | + | + | + | + | MSI-H | + | + | + | + | + | MSI-H | + | + | + | + | + | MSI-H |
| Alg-43 | + | + | + | + | + | MSI-H | + | + | + | + | - | MSI-H | + | + | + | + | - | MSI-H | + | + | + | + | - | MSI-H |
| Alg-58 | + | + | + | + | + | MSI-H | + | + | + | + | + | MSI-H | + | + | + | + | + | MSI-H | + | + | + | + | + | MSI-H |
| Alg-83 | + | + | + | + | + | MSI-H | + | + | + | + | + | MSI-H | + | + | + | + | + | MSI-H | + | + | + | + | + | MSI-H |
| Alg-89 | + | + | + | + | + | MSI-H | + | + | + | + | + | MSI-H | + | + | + | + | + | MSI-H | + | + | + | + | + | MSI-H |

| | |
|---|---|
| - | stable |
| + | unstable |
| na | failed |

**New Multiplex Droplet Digital PCR for highly sensitive detection of microsatellite instability**

**Supplementary Table 8.** Comparison of ddPCR MSI with manual thresholding for 14 FF and FFPE samples when tested at an input per well of 18, 5 and 1 ng/well, measured by Qubit dsDNA BR Assay. Seven (7) of the samples were determined as MSS and 7 as MSI-H by the Promega MSI Analysis System. The ddPCR MSI results reflect data for replicate 2 of a total of 2 replicates.

| | Promega MSI Analysis | | | | | | ddPCR MSI - 18 ng/rxn | | | | | | ddPCR MSI - 5 ng/rxn | | | | | | ddPCR MSI - 1 ng/rxn | | | | | |
|---|---|---|---|---|---|---|---|---|---|---|---|---|---|---|---|---|---|---|---|---|---|---|---|---|
| **Sample** | **BAT-25** | **BAT-26** | **NR-21** | **NR-24** | **Mono-27** | **Status** | **BAT-25** | **BAT-26** | **NR-21** | **NR-24** | **Mono-27** | **Status** | **BAT-25** | **BAT-26** | **NR-21** | **NR-24** | **Mono-27** | **Status** | **BAT-25** | **BAT-26** | **NR-21** | **NR-24** | **Mono-27** | **Status** |
| **Alg-1** | - | - | - | - | - | MSS | - | + | - | - | - | MSS | - | - | - | - | - | MSS | - | - | - | - | - | MSS |
| **Alg-2** | - | - | - | - | - | MSS | - | - | - | - | - | MSS | - | - | - | - | - | MSS | - | - | - | - | - | MSS |
| **Alg-14** | - | - | - | - | - | MSS | - | - | na | na | na | na | - | - | - | - | - | MSS | - | - | - | - | - | MSS |
| **Alg-16** | - | - | - | - | - | MSS | na | na | na | na | - | na | - | - | - | - | - | MSS | - | - | - | - | - | MSS |
| **Alg-67** | - | - | - | - | - | MSS | - | + | na | na | - | na | - | + | - | - | - | MSS | - | - | - | - | - | MSS |
| **Alg-87** | - | - | - | - | - | MSS | - | - | - | - | - | MSS | - | - | - | - | - | MSS | - | - | - | - | - | MSS |
| **Alg-100** | - | - | - | - | - | MSS | - | - | - | - | - | MSS | - | - | - | - | - | MSS | - | - | - | - | - | MSS |
| **Alg-28** | + | + | + | + | + | MSI-H | + | + | + | + | - | MSI-H | + | + | + | + | - | MSI-H | + | + | + | + | - | MSI-H |
| **Alg-30** | + | + | + | - | - | MSI-H | + | + | + | - | + | MSI-H | + | + | - | + | + | MSI-H | + | + | - | - | + | MSI-H |
| **Alg-33** | + | + | + | + | + | MSI-H | + | + | + | + | + | MSI-H | + | + | + | + | + | MSI-H | + | + | + | + | + | MSI-H |
| **Alg-43** | + | + | + | + | + | MSI-H | + | + | + | + | - | MSI-H | + | + | + | + | - | MSI-H | + | + | + | + | - | MSI-H |
| **Alg-58** | + | + | + | + | + | MSI-H | + | + | na | na | na | na | + | + | + | + | + | MSI-H | + | + | + | + | + | MSI-H |
| **Alg-83** | + | + | + | + | + | MSI-H | + | + | + | + | + | MSI-H | + | + | + | + | + | MSI-H | + | + | + | + | + | MSI-H |
| **Alg-89** | + | + | + | + | + | MSI-H | + | + | + | + | + | MSI-H | + | + | + | + | + | MSI-H | + | + | + | + | + | MSI-H |

| | |
|---|---|
| - | stable |
| + | unstable |
| na | failed |

**New Multiplex Droplet Digital PCR for highly sensitive detection of microsatellite instability**

**Supplementary Table 9. Intra- and inter-run reproducibility of sample testing.** Nine runs for cohort 1 (ALGECOLS) were analyzed for intra-and inter-run reproducibility by analyzing the positive controls run in duplicate for each run. Intra-run analysis of the positive control resulted in a mean difference in mutant fractional abundance ranging from 5.8% to 13.2% across the 5 target loci. The mean difference in wild type concentration for the 3 assays ranged from 2.8% to 3.5%. Inter-run analysis of the positive control resulted in a mutant fractional abundance CV ranging from 4.0% to 9.6% (n = 18) across the 5 target loci. The CV of the wild type concentration ranged from 2.0% to 2.5% (n = 18).

| **Marker** | **Mean Difference in Mutant Fractional Abundance between Duplicates** | **Mean Mutant Fractional Abundance (n = 18)** | **CV of Mutant Fractional Abundance (n = 18)** |
|---|---|---|---|
| BAT-25 | 8.1% | 7.1% | 5.8% |
| BAT-26 | 11.4% | 6.9% | 7.8% |
| NR-21 | 13.2% | 6.9% | 9.6% |
| NR-24 | 5.8% | 7.5% | 4.0% |
| Mono-27 | 10.6% | 6.0% | 7.9% |
| **Wild Type** | **Mean Difference in Wild Type Concentration between Duplicates** | **Mean Wild Type Concentration (copies/µL) (n = 18)** | **CV of Wild Type Concentration (n = 18)** |
| Assay 1 | 3.4% | 170.0 | 2.4% |
| Assay 2 | 3.5% | 169.5 | 2.5% |
| Assay 3 | 2.8% | 172.0 | 2.0% |

**New Multiplex Droplet Digital PCR for highly sensitive detection of microsatellite instability**

**Supplementary Table 10. Comparison of duplicate analysis.** For cohort 1, 102 samples were run in duplicate within the same run. Eighty-eight (88) of the 102 had valid results for both replicates. Among these 88 samples, the mean difference in wild type concentration for the 3 assays ranged from 4.3% to 4.5%. The marker results for these 88 samples had a 100% match between replicates for 86 of the samples. For the remaining 2 samples, detection of one marker changed from positive to negative because the mutant fractional abundance of these markers was near the limit of detection. These discordant marker results did not change the MSI status determination for these samples. Of the 88 samples, the ddPCR MSI assay determined 16 were MSI-H. The mean difference in mutant fractional abundance for these 16 samples ranged between 2.1% and 6.5% for the 5 target loci.

| | **Mean Difference between Wild Type Concentration of Duplicates** | | | **Standard Deviation of Difference between Wild Type Concentration of Duplicates** | | |
|---|---|---|---|---|---|---|
| **Wild Type** | **MSS (n = 72)** | **MSI-H (n = 16)** | **All Samples (n = 88)** | **MSS (n = 72)** | **MSI-H (n = 16)** | **All Samples (n = 88)** |
| Assay 1 | 4.2% | 4.7% | 4.3% | 5.3% | 6.2% | 5.4% |
| Assay 2 | 4.5% | 4.4% | 4.5% | 6.0% | 5.6% | 5.9% |
| Assay 3 | 4.4% | 3.8% | 4.3% | 5.7% | 4.9% | 5.5% |
| **Marker** | **Mean Difference in Mutant Fractional Abundance of MSI-H n = 16** | | | **Standard Deviation of Difference in Mutant Fractional Abundance of MSI-H n = 16** | | |
| BAT-25 | 5.5% | | | 10.0% | | |
| BAT-26 | 4.1% | | | 7.3% | | |
| NR-21 | 2.1% | | | 3.1% | | |
| NR-24 | 6.5% | | | 12.8% | | |
| Mono-27 | 5.8% | | | 11.5% | | |
